# BMI and Varus Malalignment Compound to Define a High-Risk Phenotype for Compartment-Specific Knee Osteoarthritis Progression

**DOI:** 10.64898/2026.04.15.26350819

**Authors:** McKenzie S. White, Feliks Kogan, Scott L. Delp, Constance R. Chu, Seth L. Sherman, Pai S Anoosha, Garry E. Gold, Akshay S. Chaudhari, Anthony A. Gatti

## Abstract

**Objective:** Knee osteoarthritis (KOA) is a leading cause of disability, yet which patients will experience structural decline remains unclear. Body mass index (BMI) and lower limb alignment are established risk factors for KOA, but their independent and interactive effects on compartment-specific cartilage loss and total knee replacement (TKR) have not been characterized at scale.

**Design:** We analyzed 5,930 limbs from 3,191 participants in the Osteoarthritis Initiative followed over 0-60 months. Cartilage thickness in the weight-bearing medial and lateral femur and tibia was quantified, and lower limb alignment was measured using hip-knee-ankle (HKA) angle obtained from full-limb radiographs. Linear mixed-effects models estimated the associations of BMI and lower limb alignment with longitudinal changes in cartilage thickness, modeled as interactions with time and with each other and adjusted for age, sex, and baseline Kellgren-Lawrence grade. Logistic regression with participant-clustered standard errors modeled TKR odds within five and ten years after each limb’s baseline.

**Results:** In the medial compartment, BMI and varus alignment interacted, with their combined effect exceeding the sum of independent effects (femur: β = 0.0016 mm/year per 1-SD BMI x 1-SD HKA, 95% CI [0.0001, 0.0031]; tibia: β = 0.0018, 95% CI [0.0008, 0.0028]). In the lateral compartment, alignment, but not BMI, was associated with faster cartilage loss, and its association was nonlinear (time x HKA^2^: femur β = -0.0032 mm/year, 95% CI [-0.0038, -0.0027]; tibia β = -0.0033, 95% CI [-0.0040, -0.0026]). TKR odds increased 1.11-fold at five years (95% CI [1.04, 1.18]) and 1.14-fold at ten years (95% CI [1.08, 1.21]) per 1-degree greater absolute HKA deviation from the radiographically healthy reference angle (−1.0°). BMI did not modify the association between HKA deviation and TKR.

**Conclusion:** BMI and lower limb alignment influence structural KOA progression through compartment-specific pathways. The compounding interaction in the medial compartment identifies high BMI combined with varus malalignment as a high-risk phenotype for medial structural progression.

## INTRODUCTION

Knee osteoarthritis (KOA) is a leading cause of pain and disability worldwide,[1] yet its progression is highly heterogeneous.[2] Identifying patients at greatest structural risk, those most likely to experience rapid cartilage loss and progress to total knee replacement (TKR), remains a critical challenge for clinical management and disease-modifying trial design.[3, 4] Two of the most readily measured clinical characteristics, body mass index (BMI) and lower limb alignment, are established independent risk factors for KOA,[5–9] but their combined associations with compartment-specific structural progression have not been established at scale.

While KOA is considered a heterogeneous, whole-joint disease with multiple contributing factors (e.g., inflammatory, metabolic), mechanical loading is a widely recognized driver of structural progression[10]. During gait, BMI and lower limb alignment influence joint loading through distinct pathways: body mass scales the ground reaction force and thus total joint load, while varus-valgus alignment alters how that load is distributed between the medial and lateral compartments. Consistent with this mechanical framework, both factors are independently associated with structural disease. Higher BMI has been associated with KOA incidence (BMI >27.5 kg/m², odds ratio [OR] = 3.3)[5] and progression (BMI 25–30 kg/m², relative risk = 2.4).[6] Malalignment has been associated with up to a five-fold increase in the odds of compartment-specific KOA progression.[6–9] If progression is driven by the total load experienced in a given compartment, BMI and alignment should statistically interact, producing disproportionately greater structural compartment-specific degeneration in individuals with both higher BMI and greater degrees of malalignment.

However, whether BMI and alignment interact to influence compartment-specific KOA progression remains unclear. Prior studies report conflicting results: some report primarily independent effects,[6, 11] others suggest BMI-related progression is most evident at intermediate levels of malalignment,[11] or that the cross-sectional association between BMI and OA severity is partly explained by varus malalignment,[12] and others find BMI-related progression only in neutral or valgus aligned knees.[13] These inconsistencies may partly reflect methodological limitations. Most prior work has relied on joint space narrowing (JSN) as an indirect measure of cartilage loss, rather than directly measuring cartilage thickness.[8, 9, 14, 15] Additionally, lower limb alignment has often been estimated from the femorotibial angle (FTA) on fixed-flexion knee radiographs, which correlates only moderately with full-limb hip-knee-ankle (HKA) measurements (r = 0.68-0.72).[6, 11, 16] Finally, sample sizes have generally been too small to reliably detect interaction effects.

Historically, the labor-intensive nature of quantitative cartilage assessment has impeded large-scale characterization of longitudinal cartilage thickness changes. Deep learning-based segmentation, combined with the Osteoarthritis Initiative (OAI), now enables longitudinal assessment of cartilage thickness at scale. In parallel, full-limb radiographs available in the OAI provide accurate and reproducible measures of lower limb alignment. Together, these resources enable quantification of how BMI and lower limb alignment independently and jointly associate with structural cartilage progression.

Accordingly, we quantified the independent and interactive associations of BMI and lower limb alignment with rates of compartment-specific cartilage thickness change and the risk of TKR using large-scale quantitative MRI and full-limb HKA radiographic data from the OAI. By examining these readily measured clinical factors in relation to longitudinal cartilage structural decline and TKR risk, we aimed to characterize patterns of structural progression associated with mechanical factors: BMI, malalignment, or their combination.

## METHODS

### Dataset

Data were derived from the OAI. The OAI study was approved by institutional review boards at the participating institutions, and all participants provided written informed consent. Eligible limbs had a full-limb HKA measurement, complete baseline age, sex, BMI and Kellgren-Lawrence (KL) grade, an MRI at the same visit, and at least one follow-up MRI.

Full-limb radiographs were available at OAI visits V01, V03, V05 and V06, corresponding to 12, 24, 36 and 48 months after enrollment. Each limb’s baseline was its first visit with an HKA measurement, complete covariates and an eligible MRI. Follow-up time was measured from that visit, with eligible MRIs included for up to 60 months. Additional acquisition, image and segmentation exclusions are detailed in Supplementary Methods. The final cohort included 20,843 MRI examinations from 5,930 limbs in 3,191 participants (Figure 1).

**Figure 1.**
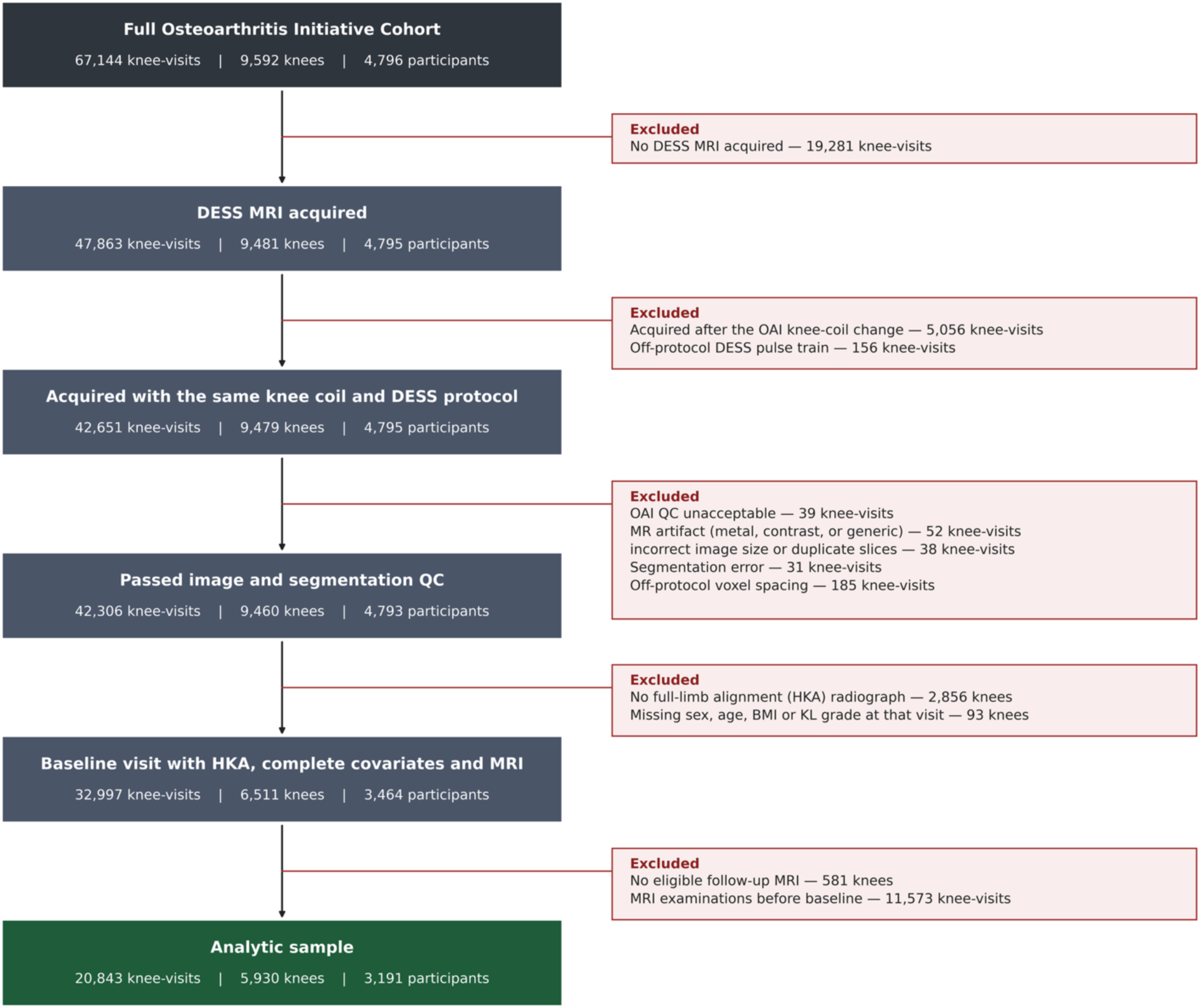
Flow diagram of knee-visit, limb (knee), and participant selection from the Osteoarthritis Initiative (OAI). The full OAI cohort comprised 67,144 potential knee-visits from 4,796 participants, two knees per participant, and seven imaging visits. Knee-visits were removed if a DESS MRI was not acquired, was obtained using an incompatible acquisition (e.g., 8-channel knee coil), or did not pass image- and segmentation-level quality control. Limbs were required to have a full-limb hip-knee-ankle (HKA) alignment radiograph, complete baseline covariates (sex, age, BMI, and Kellgren-Lawrence grade), and an MRI at that same baseline visit, together with at least one additional MRI examination after baseline. MRI examinations obtained before the eligible baseline visit were excluded. Each process box reports the number of knee-visits, limbs, and participants remaining; the boxes to the right list the exclusions applied at each step. The final analytic sample comprised 20,843 MRI examinations from 5,930 limbs of 3,191 participants.

### Lower Limb Alignment

HKA was measured semiautomatically from full-limb radiographs by OAI Projects 32 and 60. In a validation study, the method used by Project 60 had HKA inter-reader reliability intraclass correlation coefficient [ICC] = 0.995 and intra-reader ICC = 0.998.[17] Project 32 measurements agreed closely with Project 60 (ICC = 0.98; mean difference = 0.1°). When both projects provided a measurement, we used their mean; otherwise, we used the available measurement. Negative HKA values indicate varus alignment, and positive values indicate valgus alignment.

### Cartilage Thickness Measurements

Femoral and tibial bone and cartilage were segmented from sagittal Double Echo in Steady State MRI using validated deep-learning methods[18] and surface meshes were generated.[19] Standardized regions of interest were defined on reference meshes to include full-thickness cartilage lesions (0 mm).[20, 21] Cartilage thickness was measured between the bone-cartilage interface and articular surface and averaged within the weight-bearing medial and lateral femur and medial and lateral tibia. Methodological details are provided in Supplementary Methods, and comparisons to manual thickness measurements are presented in Supplementary Table 1. Notably, the four OAI MRI scanners underwent a platform upgrade (Supplementary Figure 1) associated with a systematic decrease in measured cartilage thickness beyond typical inter-visit change (Supplementary Figures 2-3). We corrected post-upgrade thickness values using scanner-and region-specific offsets (Supplementary Table 2). Supplementary Tables 4A–B show results using uncorrected data.

### Statistical Analyses

#### Modeling the Associations of BMI and Alignment with the Rate of Cartilage Thinning

To evaluate the independent and interactive effects of BMI and lower limb alignment on changes in longitudinal cartilage thickness, we fitted separate linear mixed-effects models for the medial femur, medial tibia, lateral femur, and lateral tibia. The outcome was cartilage thickness measured repeatedly at baseline and follow-up visits. Baseline thickness entered the primary model as the outcome at time zero rather than as a separate covariate. Elapsed time was modeled continuously in years from each limb’s baseline and centered at the mean observed follow-up time. The time coefficient estimates average annual change in cartilage thickness (mm/year) at the mean BMI and HKA alignment. Interactions with time estimate how this annual rate differs as a function of BMI and HKA alignment.

We tested the time x HKA x BMI interaction in all four compartments and omitted it when nonsignificant. All models retained the main effects and two-way interactions among time, HKA, and BMI. Lateral models also included HKA^2^ and time x HKA^2^ to model nonlinear associations with cartilage thickness and its annual rate of change (Supplementary Tables 3B, 6).

All primary cartilage models were adjusted for baseline age, sex, and categorical KL grade. BMI, HKA, and age were z-score normalized [(X – mean)/standard deviation] to improve model convergence. Cartilage thickness remained in millimeters. Model coefficients are reported using standardized predictors, whereas clinical examples and predictions use BMI in kg/m² and HKA in degrees. Fitted models predicted mean cartilage thickness at baseline and five years (Supplementary Tables 5A–D).

Models included a participant-level random intercept and a limb-within-participant random intercept and slope for time. Models were fitted using restricted maximum likelihood, with Wald 95% confidence intervals and Satterthwaite tests for fixed effects. A Benjamini-Hochberg false discovery rate correction was applied across the four primary interaction tests (Supplementary Table 6).

### Model assumptions and sensitivity analyses

We assessed model fit using linear and nonlinear time, BMI and HKA factors with rates calculated separately for each limb (Supplementary Tables 7-8 and Supplementary Figures 4A– C). Model assumptions were assessed using residual-versus-fitted plots, residual distributions, and quantile-quantile plots of residuals and random effects (Supplementary Figures 5A–D).

Sensitivity analyses evaluated alternative baseline severity measures, restrictions to limbs without regional baseline JSN or with baseline KL grade 0, visit-specific and average BMI over the study protocol, physical activity adjustment, and body mass replacing BMI (Supplementary Tables 9–12).

### Modeling the Association of BMI and Alignment with Total Knee Replacement

To evaluate how BMI and HKA alignment were associated with total knee replacement (TKR), we used logistic regression for confirmed TKR within five years and within ten years of each limb’s baseline. Limbs with unconfirmed replacement reports were excluded, as were limbs without recorded TKR whose follow-up ended before the relevant cutoff. Absolute HKA deviation was calculated from the mean alignment of a radiographically healthy reference subgroup with KL grade 0 at baseline and every matched follow-up assessment (mean HKA = −1.0°). We first fitted models including absolute HKA deviation, BMI, and their interaction, adjusted for baseline age, sex, and KL grade. Standard errors were clustered by participant to account for correlation between limbs. When the interaction was nonsignificant, it was removed and the model was refit with absolute HKA deviation, BMI, and all adjustment variables to estimate adjusted odds ratios with 95% confidence intervals. Further details on TKR follow-up and modeling are provided in Supplementary Methods.

### Data and Code Availability

Data generated in this study, including quantitative cartilage thickness, statistical analysis scripts, and model outputs, are publicly available at https://github.com/kenziew/oai_hka_bmi.

## RESULTS

### Dataset

The analytic cohort comprised 20,843 MRI examinations from 5,930 limbs of 3,191 participants, of whom 2,739 (85.8%) contributed both limbs (Figure 1). Mean age was 62.9 ± 9.1 years, BMI was 28.7 ± 4.8 kg/m², and HKA was −1.3° ± 3.4° (Table 1). The distribution of baseline visits relative to OAI enrollment is shown in Supplementary Table 13. Confirmed TKR occurred in 190/5,507 limbs (3.5%) with observed five-year status and 405/4,155 limbs (9.7%) with observed ten-year status.

**Table 1:** Characteristics of the Study Cohort and Distribution of BMI and Alignment.

|  | Limbs (n) | Participants (n) | Age (years) | BMI (kg/m<sup>2</sup>) |
| --- | --- | --- | --- | --- |
| **Total Sample** | 5930 | 3191 | 62.9 ± 9.1 | 28.7 ± 4.8 |
| Female |  | 1797 (56.3 %) | 63.3 ± 8.9 | 28.6 ± 5.2 |
| Male |  | 1394 (43.7 %) | 62.4 ± 9.3 | 28.7 ± 4.1 |
| **Number of time points contributed, including baseline** |  |  |  |  |
|  | Limbs(n) | Participants (n) |  |  |
| 2 | 1021 | 457 |  |  |
| 3 | 1942 | 937 |  |  |
| 4 | 1860 | 1046 |  |  |
| 5 | 1107 | 751 |  |  |
| **MRI availability at each time point** |  |  |  |  |
|  | Limbs(n) | Participants (n) |  |  |
| Baseline | 5930 | 3191 |  |  |
| 1 year | 5019 | 2930 |  |  |
| 2 years | 3738 | 2199 |  |  |
| 3 years | 3017 | 1681 |  |  |
| 4 years | 1576 | 815 |  |  |
| 5 years | 1563 | 848 |  |  |
| **Descriptive Body Mass Index and Alignment Categories at Baseline Per Limb** |  |  |  |  |
| | Underweight<br>($<18.5$ kg/m<sup>2</sup>) | Healthy<br>($18.5$-$24.9$ kg/m<sup>2</sup>) | Overweight<br>($25.0$-$29.9$ kg/m<sup>2</sup>) | Obese<br>($\geq 30$ kg/m<sup>2</sup>) |
| Severe Varus<br>$< -8.8^\circ$ | 0 | 17 | 40 | 45 |
| Moderate Varus<br>$\geq -8.8^\circ$ to $< -6.2^\circ$ | 0 | 49 | 113 | 138 |
| Mild Varus<br>$\geq -6.2^\circ$ to $< -3.6^\circ$ | 1 | 236 | 391 | 325 |
| Neutral<br>$\geq -3.6^\circ$ to $\leq 1.6^\circ$ | 11 | 837 | 1383 | 1266 |
| Mild Valgus<br>$> 1.6^\circ$ to $\leq 4.2^\circ$ | 3 | 200 | 315 | 295 |
| Moderate Valgus<br>$> 4.2^\circ$ to $\leq 6.8^\circ$ | 1 | 41 | 72 | 76 |
| Severe Valgus<br>$> 6.8^\circ$ | 0 | 14 | 34 | 27 |

Alignment categories were defined relative to a radiographically healthy reference subgroup with KL grade 0 at baseline and at all observed follow-up assessments (n = 1,728 limbs from 1,197 participants; mean HKA = $-1.0^\circ \pm 2.6^\circ$ ). Limbs with a missing KL grade at any included visit were excluded from the reference subgroup. Neutral = $\pm 1$ SD ( $-3.6^\circ$ to $1.6^\circ$ ); mild = 1–2 SD; moderate = 2–3 SD; severe = $> 3$ SD.

The radiographically healthy reference subgroup included 1,728 limbs from 1,197 participants with KL grade 0 at baseline and every observed follow-up visit. Mean age was 61.5 ± 9.2 years, BMI was 27.4 ± 4.4 kg/m², HKA was −1.0° ± 2.6°, and 51.0% were female. Table 1 summarizes WHO BMI categories and alignment categories defined relative to this subgroup’s HKA distribution. These categories were descriptive; BMI and HKA were modeled continuously.

### Influence of BMI and Lower Limb Alignment on the Rate of Cartilage Thinning Medial Compartment

Baseline cartilage thickness and radiographic joint-space narrowing grades by BMI and alignment category are reported in Supplementary Tables 14A–B. At cohort-average BMI and alignment, medial femoral thinning was 0.0191 mm/year (95% CI [0.0177, 0.0205]; 0.99% of mean baseline thickness). Higher BMI and greater varus alignment were jointly associated with faster thinning beyond their additive contributions (time x BMI x HKA: β = 0.0016 mm/year per 1-SD BMI x 1-SD HKA, 95% CI [0.0001, 0.0031], p = 0.042; Figures 2A and 3A; Supplementary Table 3A). Predicted cartilage thickness trajectories over five years illustrate how the combination of higher BMI and varus alignment produces greater thinning than either factor alone (Figure 3A). Each additional 1 kg/m² of BMI at mean HKA was associated with 7.6% faster thinning, and each additional 1° of varus at mean BMI with 20.5% faster thinning (Supplementary Figure 6A). For combined increases of 1 kg/m² and 1° of varus, the interaction contributed an additional 0.51% of the thinning rate at mean BMI and HKA. At increases of 5 kg/m² and 5° of varus, total predicted thinning was 153.6% faster (0.0484 mm/year; Table 2).

**Figure 2.**
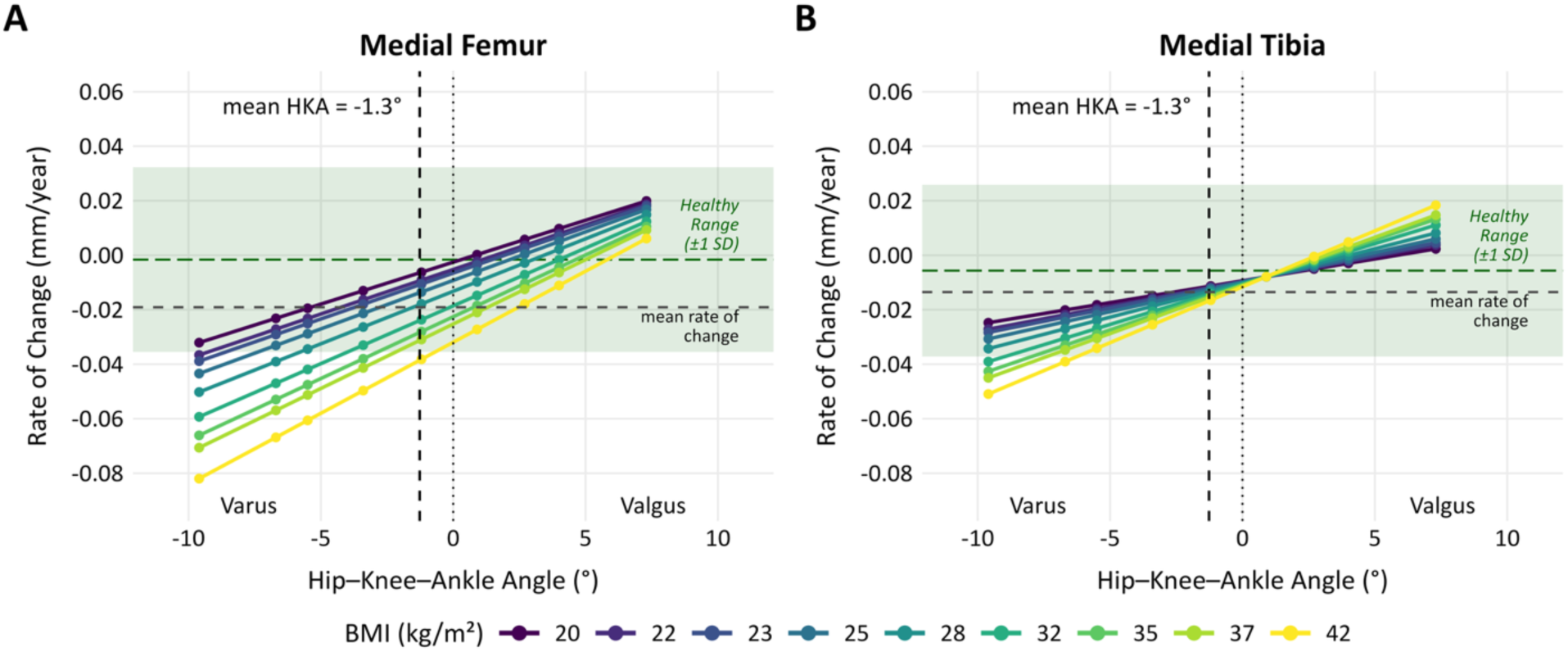
Predicted Annual Rate of Cartilage Thinning in the Medial Compartments. Predicted rate of thinning (mm/year) is shown across Hip-Knee-Ankle (HKA) alignment for the medial femur (A) and medial tibia (B). Colored lines represent BMI at the 1st, 5th, 10th, 25th, 50th, 75th, 90th, 95th, and 99th participant-level percentiles (20–42 kg/m²); dots mark predictions at the corresponding limb-level HKA percentiles (-9.6° to 7.3°). Negative rates indicate thinning. A significant three-way interaction was observed between time, BMI, and HKA angle (medial femur: p = 0.042; medial tibia: p < 0.001), indicating that higher BMI and greater varus alignment were jointly associated with faster thinning beyond their additive contributions. Black vertical dashed lines indicate mean HKA (-1.3°). Horizontal grey dashed lines show predicted annual change at mean BMI and HKA (medial femur: -0.0191 mm/year; medial tibia: -0.0135 mm/year). Green dashed lines show model-estimated rates at the same BMI and HKA reference values in the radiographically healthy subgroup (1,728 limbs from 1,197 participants with KL grade 0 at baseline and throughout observed follow-up). Green bands extend one SD of the subgroup’s observed limb-level annual change rates above and below these reference rates.

**Figure 3.**
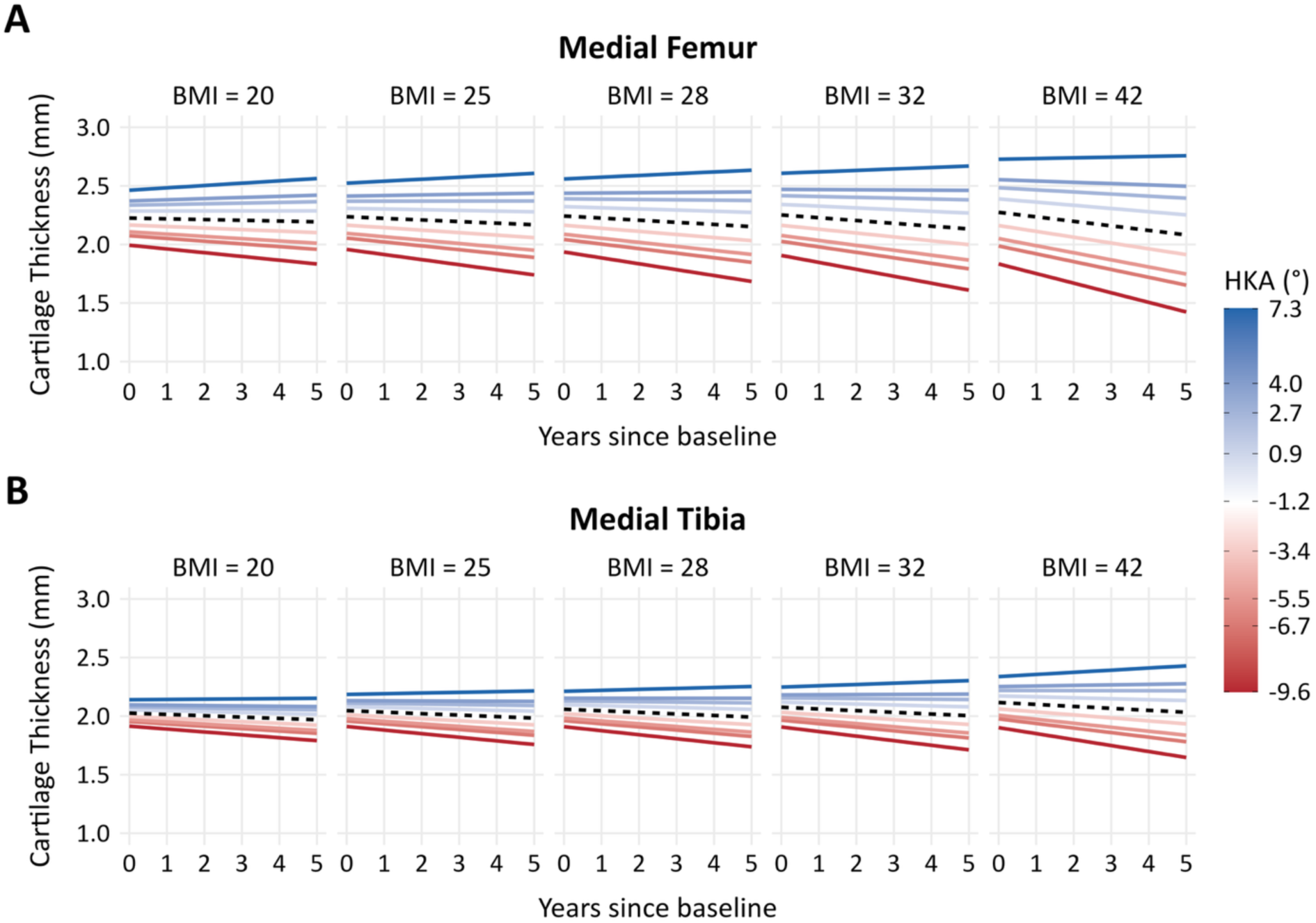
Predicted Longitudinal Cartilage Thickness Trajectories for the Medial Femur (A) and Medial Tibia (B). Models show predicted cartilage thickness over five years after limb baseline. Columns represent BMI at the 1st, 25th, 50th, 75th, and 99th participant-level percentiles (20, 25, 28, 32, and 42 kg/m²). Colored lines represent HKA at the 1st, 5th, 10th, 25th, 50th, 75th, 90th, 95th, and 99th limb-level percentiles (-9.6° to 7.3°). Predictions are shown at mean age, male sex, and baseline KL grade 0. The black dashed line in each panel shows the trajectory at mean HKA (-1.3°) for that panel’s BMI. While divergence between varus (red) and valgus (blue) alignment is present at all BMIs, this gap progressively widens as BMI increases, demonstrating the compounding effect of BMI and varus alignment on medial cartilage thinning.

**Table 2.** Model-Predicted Compounding Associations of BMI and Varus Alignment with Medial Cartilage Thinning. Each unit represents simultaneous increases of 1 kg/m² in BMI and 1° of varus from mean BMI (28.7 kg/m²) and HKA (-1.3°). Percentages express the BMI, alignment, and interaction contributions on a common scale relative to the reference thinning rate; positive values indicate faster thinning. The interaction is the additional contribution beyond BMI and alignment alone. Negative annual thickness changes indicate thinning; brackets show 95% confidence intervals.

|  | Increase level | BMI Effect (%) | Varus Effect (%) | Interaction Effect (%) | Total Increase (%) | Annual Thickness Change (mm/year) [95% CI] |
| --- | --- | --- | --- | --- | --- | --- |
| Reference | - | - | - | - | - | -0.0191 [-0.0205, -0.0177] |
| Medial Femur | +1 Unit | 7.6 % | 20.5 % | 0.5 % | 28.7 % | -0.0245 [-0.0261, -0.0230] |
|  | +2 Unit | 15.3 % | 41.1 % | 2.0 % | 58.4 % | -0.0302 [-0.0320, -0.0284] |
|  | +5 Unit | 38.2 % | 102.7 % | 12.7% | 153.6 % | -0.0484 [-0.0520, -0.0447] |
| Reference | - | - | - | - | - | -0.0135 [-0.0144, -0.0126] |
| Medial Tibia | +1 Unit | 1.8 % | 19.1 % | 0.8 % | 21.8% | -0.0165 [-0.0174, -0.0155] |
|  | +2 Unit | 3.6 % | 38.3 % | 3.4 % | 45.3 % | -0.0196 [-0.0208, -0.0185] |
|  | +5 Unit | 9.0 % | 95.7 % | 21.0 % | 125.8 % | -0.0305 [-0.0329, -0.0281] |

At the cohort-average BMI and alignment, medial tibial thinning was 0.0135 mm/year (95% CI [0.0126, 0.0144]; 0.71% of mean baseline thickness; Figure 2B). The time x BMI x HKA interaction was significant (β = 0.0018 mm/year per 1-SD BMI x 1-SD HKA, 95% CI [0.0008, 0.0028], p < 0.001; Figures 2B and 3B; Supplementary Table 3A). Each +1 kg/m² in BMI at mean HKA was associated with 1.8% faster thinning, and each additional 1° of varus at mean BMI with 19.1% faster thinning (Supplementary Figure 6B). For combined increases of 1 kg/m² and 1° of varus, the interaction contributed an additional 0.84% of the thinning rate at mean BMI and HKA. At increases of 5 kg/m² and 5° of varus, total predicted thinning was 125.8% faster (0.0305 mm/year; Table 2).

### Lateral Compartment

Neither lateral compartment showed a significant time x BMI x HKA interaction in the initial models (femur: p = 0.413; tibia: p = 0.250). The time x HKA² term was significant in both compartments (femur: β = −0.0032, 95% CI [−0.0038, −0.0027]; tibia: β = −0.0033, 95% CI [−0.0040, −0.0026]; both p < 0.001; Supplementary Table 3B), with thinning slowest at approximately 3° of varus in the lateral femur and 5° of varus in the lateral tibia, and faster toward both extremes (Supplementary Figure 6C–D). BMI was not significantly associated with lateral thinning (femur: p = 0.242; tibia: p = 0.757).

### Influence of BMI and Lower Limb Alignment on the Risk of Total Knee Replacement

The BMI x absolute HKA deviation interaction was not significant at five years (β = −0.0487, 95% CI [−0.1533, 0.0559], p = 0.361) or ten years (p = 0.286). In the models without the interaction, each additional degree of deviation from reference HKA (−1.0°) was associated with higher TKR odds at five years (OR = 1.11, 95% CI [1.04, 1.18], p = 0.001) and ten years (OR = 1.14, 95% CI [1.08, 1.21], p < 0.001; Figure 4). A five-degree deviation corresponded to 1.67 times the odds of TKR at five years (95% CI [1.22, 2.29]) and 1.96 times the odds at ten years (95% CI [1.50, 2.55]; Figure 4A–B). BMI was not significantly associated with TKR odds at five years (OR = 1.03 per kg/m², 95% CI [0.99, 1.06], p = 0.126) or ten years (OR = 1.02 per kg/m², 95% CI [0.99, 1.05], p = 0.111; Supplementary Table 15).

**Figure 4.**
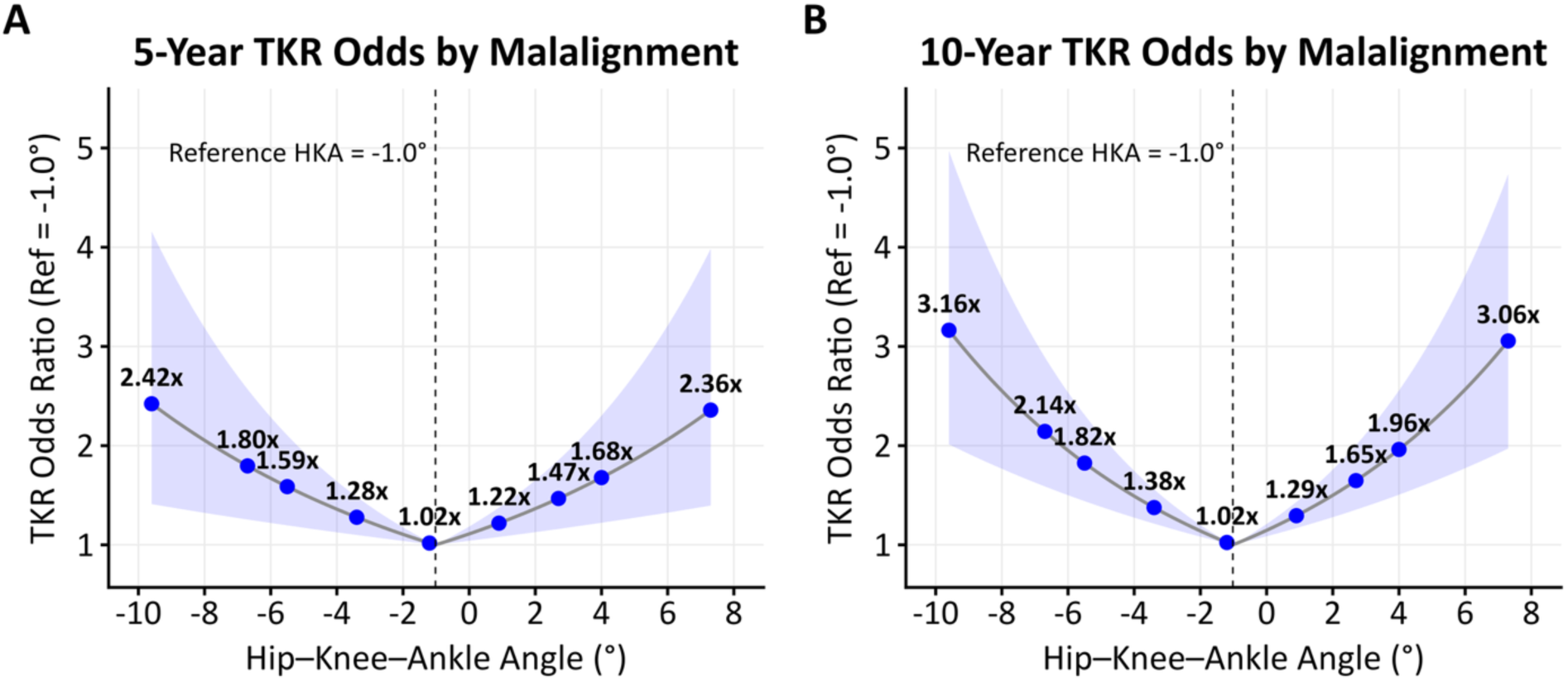
Five- and Ten-Year Total Knee Replacement Odds by Baseline Alignment. Adjusted odds ratios are relative to the mean HKA of the radiographically healthy subgroup (-1.0°), from the five-year (A) and ten-year (B) main-effects logistic models adjusted for BMI, age, sex, and baseline KL. Follow-up windows were measured from each limb’s baseline. Blue dots represent full-cohort baseline HKA 1st, 5th, 10th, 25th, 50th, 75th, 90th, 95th, and 99th percentiles (-9.6° to 7.3°), and shading represents pointwise 95% confidence intervals using participant-clustered robust standard errors. The symmetric curves follow the absolute-deviation model specification. Each additional degree of HKA deviation was associated with an odds ratio of 1.11 (95% CI 1.04– 1.18) at five years and 1.14 (95% CI 1.08–1.21) at ten years.

### Sensitivity Analyses

The medial time x BMI x HKA and lateral time x HKA² interactions persisted when models used BMI measured at each MRI visit or mean BMI across follow-up, rather than baseline BMI carried forward (Supplementary Table 10), with physical activity adjustment (Supplementary Table 11), and when baseline cartilage thickness or JSN replaced KL grade (Supplementary Tables 9A–B). In the quantitative joint space width subset, both medial interactions weakened and became nonsignificant with either baseline KL or quantitative joint space width adjustment (Supplementary Table 9C). Among limbs without baseline JSN, the medial femoral time x BMI x HKA and lateral femoral time x HKA² interactions weakened and were no longer significant (Supplementary Table 9D). When modeling only limbs with baseline KL grade 0, both medial interactions persisted. Lateral femoral time x HKA² was nonsignificant, whereas lateral tibial time x HKA² remained significant (p = 0.004; Supplementary Table 9E). Replacing BMI with body mass retained the medial interactions and lateral time x HKA² associations (Supplementary Tables 12A–B).

## DISCUSSION

In this large-scale longitudinal study of 3,191 participants (5,930 limbs), BMI and lower limb alignment were associated with structural KOA progression in patterns that were compartment specific. In the medial compartment, BMI and varus alignment interacted, such that their combined effect on the rate of cartilage thinning exceeded the sum of their independent contributions. In the lateral compartment, cartilage thinning was faster at greater degrees of both varus and valgus alignments, while BMI showed no significant association. Finally, higher odds of TKR were associated with the degree of malalignment, while BMI showed no significant association. Together, these findings demonstrate that the relationships between BMI, lower limb alignment, and structural KOA progression are compartment-specific, and identify a high-risk phenotype for medial OA progression.

The compounding interaction observed in the medial compartment is consistent with the mechanical framework in which BMI and lower limb alignment function as compounding determinants of joint loading.[22] During gait the medial side generally carries a greater share of total joint load.[23] Higher BMI increases the load the medial compartment already bears disproportionately, while varus alignment concentrates that load further onto the same compartment.[24] Together, these factors may impose greater mechanical demands on medial cartilage than either factor alone. This biomechanical framework is consistent with gait studies linking the knee adduction moment (KAM), a widely used surrogate of medial relative to lateral compartment loading, to medial cartilage loss[25, 26] and with findings that KAM interacts with BMI to predict medial tibial cartilage volume loss over 2.5 years.[27] Our findings extend this by showing that higher BMI and varus alignment measured using full-limb HKA are jointly associated with faster medial cartilage thinning. Moyer et al. also assessed quantitative cartilage thickness using OAI data but used radiographic FTA, which only moderately correlates with full limb HKA measurements (r=0.72)[16]. Although Moyer et al. did not detect a significant BMI x alignment interaction, alignment was associated with both medial and lateral cartilage loss, whereas a significant BMI association was observed only for medial cartilage loss in knees with minor varus alignment.[11] Using FTA as a surrogate for HKA could reduce sensitivity to a BMI x alignment interaction. The shorter two-year follow-up and restriction to knees with established radiographic OA by Moyer et al. could further limit its detection.[11] Our larger cohort (5,930 limbs vs. 558 limbs), full-limb HKA measurements, and follow-up of up to five years across the disease spectrum help explain these contrasting findings.

Across prior studies, alignment has been shown to modulate the effects of BMI on radiographic KOA,[6, 12, 13, 28] with findings differing by baseline disease stage. Niu et al. found that obesity increased radiographic JSN progression in neutral but not varus knees with established OA at baseline.[13] In knees without established OA, obesity increased the risk of developing OA (KL grade ≥2) across alignment groups.[13] JSN and KL grade are coarse radiographic endpoints, while categorizing BMI and alignment can reduce statistical power. Using continuous cartilage thickness across the full disease spectrum (continuous BMI and HKA) overcome these limitations. Replacing baseline KL grade with cartilage thickness or compartment-specific JSN produced similar estimates (Supplementary Tables 9A–B). Both medial interactions also remained significant in limbs with baseline KL grade 0, supporting a compounding association even without baseline radiographic OA (Supplementary Table 9E).

Differences in how load is distributed medially and laterally may contribute to the compartment-specific BMI findings. In contrast to the medial compartment, BMI was not significantly associated with lateral cartilage thinning, either independently or through an interaction with alignment. However, substituting body mass for BMI revealed a significant association between body mass and lateral thinning, while retaining the medial interactions and lateral alignment relationships (Supplementary Tables 12A–B). The absence of a significant lateral BMI association does not rule out a contribution of body mass to lateral cartilage loss. Alignment associations differed across compartments. Whereas the primary models described a linear association between alignment and medial thinning, the association with lateral thinning was nonlinear, with faster thinning rates towards the extremes of both valgus and varus alignments (Supplementary Figure 6C–D). Because varus alignment is expected to offload the lateral compartment, faster lateral thinning in varus limbs may reflect broader joint involvement as medial disease progresses.

The differing medial and lateral findings raise a question about what constitutes a “neutral” alignment for cartilage preservation. The compartment-specific relationships suggest that alignment associated with slower thinning differs between compartments, and alignment effects on cartilage should not be assumed to operate symmetrically from 0°. Limbs that remained radiographically healthy throughout follow-up (KL grade 0 at all timepoints) had an average HKA angle of −1.0°, indicating a slight varus alignment. This slight varus position is consistent with radiographic health but does not identify the alignment most favorable for cartilage preservation. Identifying such an alignment would help inform surgical planning and requires evidence from intervention studies.

In the TKR analysis, greater absolute deviation from the reference subgroup mean alignment (-1.0°) was associated with higher five-year TKR odds, with similar findings at ten years (Supplementary Table 15). This model assumes equivalent associations for varus and valgus deviations from the mean. BMI, however, was not significantly associated with TKR odds despite its association with faster medial cartilage thinning. The dissociation between BMI’s role in increasing the rate of cartilage thinning and its lack of association with TKR risk appears counterintuitive but may reflect systematic biases inherent to the surgical selection process.[29] Many centers apply upper BMI thresholds for TKR eligibility due to concerns about perioperative complications and inferior outcomes in patients with obesity,[29] which would weaken the observed association between BMI and TKR. Furthermore, TKR can also correct for varus or valgus malalignment,[30, 31] potentially favoring patients with deformity and strengthening the alignment–TKR association. Further work is needed to distinguish the influence of surgical selection criteria from underlying structural disease progression in determining which patients progress to TKR. These findings have implications for disease-modifying OA drug (DMOAD) trial endpoint development. Several proposed intermediate endpoints for clinical trials, including end-stage KOA (esKOA) and the Composite Knee Osteoarthritis Symptom Outcome (CKOASO), were developed or validated against subsequent TKR as a measure of clinical relevance.[32–34] If access to TKR is systematically less likely in high-BMI patients regardless of structural severity, these endpoints may inherit the same surgical selection bias, potentially underweighting BMI as a structural risk factor and miscalibrating progression thresholds for patients at substantial risk of structural deterioration. BMI and alignment should therefore be considered in both enrollment and endpoint validation in disease-modifying trials.

Our compartment-specific cartilage findings have implications for risk-factor modification in KOA. The association of higher BMI with faster medial thinning supports testing weight-loss strategies[35], including lifestyle interventions targeting diet and exercise[36] and, where appropriate, glucagon-like peptide-1 (GLP-1) based pharmacotherapies,[37, 38] as potential structural risk-modifying approaches for KOA. A recent GLP-1 trial demonstrated weight loss and pain benefits, but whether these benefits extend to cartilage preservation remains to be established.[37] Alignment-directed approaches may also be relevant. Changes in alignment after surgical realignment have been associated with changes in frontal-plane knee moments during gait.[39] Individuals with varus malalignment may also benefit from conservative approaches such as valgus knee bracing to reduce medial loading[40, 41] and improve pain and function.[42] In patients with substantial malalignment or failure of conservative management, particularly younger or more active individuals who are not yet candidates for arthroplasty, surgical realignment procedures such as medial opening wedge high tibial osteotomy (HTO) may be appropriate.[43] A recent randomized controlled trial demonstrated that HTO slowed medial compartment cartilage loss in patients with varus-aligned KOA, showing that mechanical realignment can modify structural disease trajectories.[44] This supports alignment as a modifiable mechanical target, consistent with the compartment-specific findings. Individuals with both elevated BMI and pronounced varus alignment may represent a group at increased risk of medial cartilage thinning and therefore, combined approaches targeting both BMI and alignment may be needed. Notably, 508 limbs (8.6%) in our cohort presented with both obesity and varus alignment (Table 1). Dedicated clinical trials testing whether interventions targeting BMI, alignment, or both slow the rate of cartilage loss and reduce the risk of TKR are needed to translate these observational findings into evidence-based treatment strategies.

Several limitations warrant consideration. The HKA angle is a frontal-plane metric that does not capture the full complexity of lower limb alignment, including the origin of deformity (femur vs. tibia), rotational alignment, or sagittal plane deformities. HKA measurements were obtained at a single baseline timepoint and reflect static alignment only; the dynamics of malalignment during gait further modulate compartment-specific loading and should be considered in future analyses. BMI and self-reported activity also incompletely characterize mechanical loading. Although our analysis focused on the mechanical pathway, we did not assess biological contributors to progression, including local inflammation[45, 46] and metabolic factors associated with elevated BMI. These mechanisms may operate alongside the loading effects described here and warrant investigation in future work. We did not assess changes in pain or function. Finally, the associations reported here are observational. Evaluation of the effects of intervention-induced changes in BMI and alignment on cartilage loss and TKR risk is needed to establish these risk factors as modifiable targets. To support reproducibility and future investigation, all quantitative cartilage thickness outcomes, analysis scripts, and model outputs are publicly available at https://github.com/kenziew/oai_hka_bmi.

This large-scale longitudinal study demonstrates that BMI and lower limb alignment interact to increase the rate of medial compartment cartilage thinning, whereas lateral cartilage loss and TKR odds were associated with alignment without significant BMI associations. These findings identify patients with elevated BMI and varus malalignment as a high-risk phenotype for structural progression and highlight both factors as candidate interventional targets for structural joint preservation.

## Data Availability

Data generated in this study, statistical analysis scripts, and model outputs will be publicly available at https://github.com/kenziew/oai_hka_bmi, which will be updated upon manuscript acceptance.

## Author Contributions

(1) Substantial contributions to the conception or design of the work; or the acquisition, analysis, or interpretation of data for the work: [MSW, AAG]
(2) Drafting the work or reviewing it critically for important intellectual content: [MSW, AAG]
(3) Final approval of the version to be published: [MSW, FK, SLD, CRC, SLS, APS, GEG, ASC, AAG]
(4) Agreement to be accountable for all aspects of the work in ensuring that questions related to the accuracy or integrity of any part of the work are appropriately investigated and resolved. [MSW, FK, SLD, CRC, SLS, APS, GEG, ASC, AAG]

### Funding Sources

1) Wu Tsai Human Performance Alliance
2) NIH R01AR079431 (Feliks Kogan -National Institute of Arthritis and Musculoskeletal and Skin Diseases)
3) R01AR083018 (Feliks Kogan -National Institute of Arthritis and Musculoskeletal and Skin Diseases)
4) R01AR077604 (Brian A. Hargreaves -National Institute of Arthritis and Musculoskeletal and Skin Diseases)
5) R01EB002524 (Garry E. Gold -National Institute of Biomedical Imaging and Bioengineering)
6) P41EB027060 (Scott L. Delp -National Institute of Biomedical Imaging and Bioengineering)
7) P50HD118632 (Scott L. Delp -Eunice Kennedy Shriver National Institute of Child Health and Human Development)
8) W81XWH-18-1-0590 (Constance R. Chu -Department of Defense)
9) I01CX002527 (Constance R. Chu -Department of Veterans Affairs)

## Declaration of generative AI and AI-assisted technologies in the manuscript preparation process

During the preparation of this work the author(s) used Claude (Anthropic) and Codex (OpenAI) to improve clarity and readability of the manuscript. After using this tool/service, the author(s) reviewed and edited the content as needed and take(s) full responsibility for the content of the published article.

