## Supplementary Material for "BMI and Varus Malalignment Compound to Define a High-Risk Phenotype for Compartment-Specific Knee Osteoarthritis Progression"

### Image and Segmentation Quality Control

The MRI protocol changed from a single-channel quadrature extremity coil to an 8-channel knee coil, primarily at OAI V10 visit. Evaluation of these data identified that the 8-channel knee coil systematically changed knee MRI volume contrast and thus cartilage thickness estimation. We validated this systematic difference in multiple deep learning-based segmentation models from UCSF, NYU, and the present dataset. Therefore, we excluded V10 examinations and any MRI acquired with the 8-channel coil. After examinations with incompatible acquisitions (e.g., coil or pulse-train changes) were removed, each remaining examination underwent two sequential quality-control stages before cohort selection. Image-level quality control used the OAI central MRI quality assessments and expert review. We excluded examinations rated unacceptable (not readable or no result), affected by an MRI artifact (metal, contrast, or other), or acquired with an incorrect image size or duplicate slices. Segmentation and protocol quality control then identified nonphysiological cartilage segmentations and incompatible voxel spacing. We excluded examinations with errors in the analyzed femoral or tibial cartilage, including absent, one-sided, or grossly asymmetric cartilage or structures fused across the intercondylar notch, and examinations with off-protocol voxel spacing. In total, 129 examinations were excluded at image-level quality control and 216 at segmentation and protocol quality control, leaving 42,306 examinations from 9,460 limbs eligible for cohort selection.

### Cartilage Thickness Measurements Quality Control

Four implausible region-level mean thickness measurements (zero thickness, or thickness below 0.5 mm in limbs with KL grade 0 or 1 at that visit) were set to missing. Two additional cartilage measurements could not be computed, one due to an incomplete segmentation and one due to a poor neural shape model fit. In total, 6 of 83,372 Region-level cartilage thickness measurements were missing. No cartilage thickness measurements were imputed or interpolated.

### Segmentation Performance and Reference-Mesh Construction

In a previously published study, the segmentation methods achieved tibiofemoral cartilage Dice similarity coefficients of 0.876–0.913 and average symmetric surface distances of 0.13-0.17 mm.[1] Average reference meshes for the femur and tibia were generated using a neural shape model previously trained on 6,325 knees from the OAI baseline visit. The reference cohort comprised knees with MRI Osteoarthritis Knee Score (MOAKS) cartilage morphology scores of 0. Standardized regions of interest followed established standards[2] and included vertices where ≥95% of this reference cohort had overlying cartilage. These common regions allowed full-thickness cartilage lesions to contribute measurements of 0 mm.

### TKR Follow-up

TKR events required confirmation of total knee replacement in the OAI outcomes dataset. Follow-up was measured from the clinical visit corresponding to each limb’s baseline. We used the latest available clinical visit date to determine follow-up, extended by any later confirmed total or partial replacement. Follow-up ended at the earliest of confirmed TKR, the last available follow-up date, death, confirmed partial replacement, or 16 years after OAI enrollment. Confirmed TKR within the five- or ten-year window was counted as an event. Limbs followed through the cutoff without TKR were counted as non-events. Limbs whose follow-up ended before the cutoff without TKR were excluded.

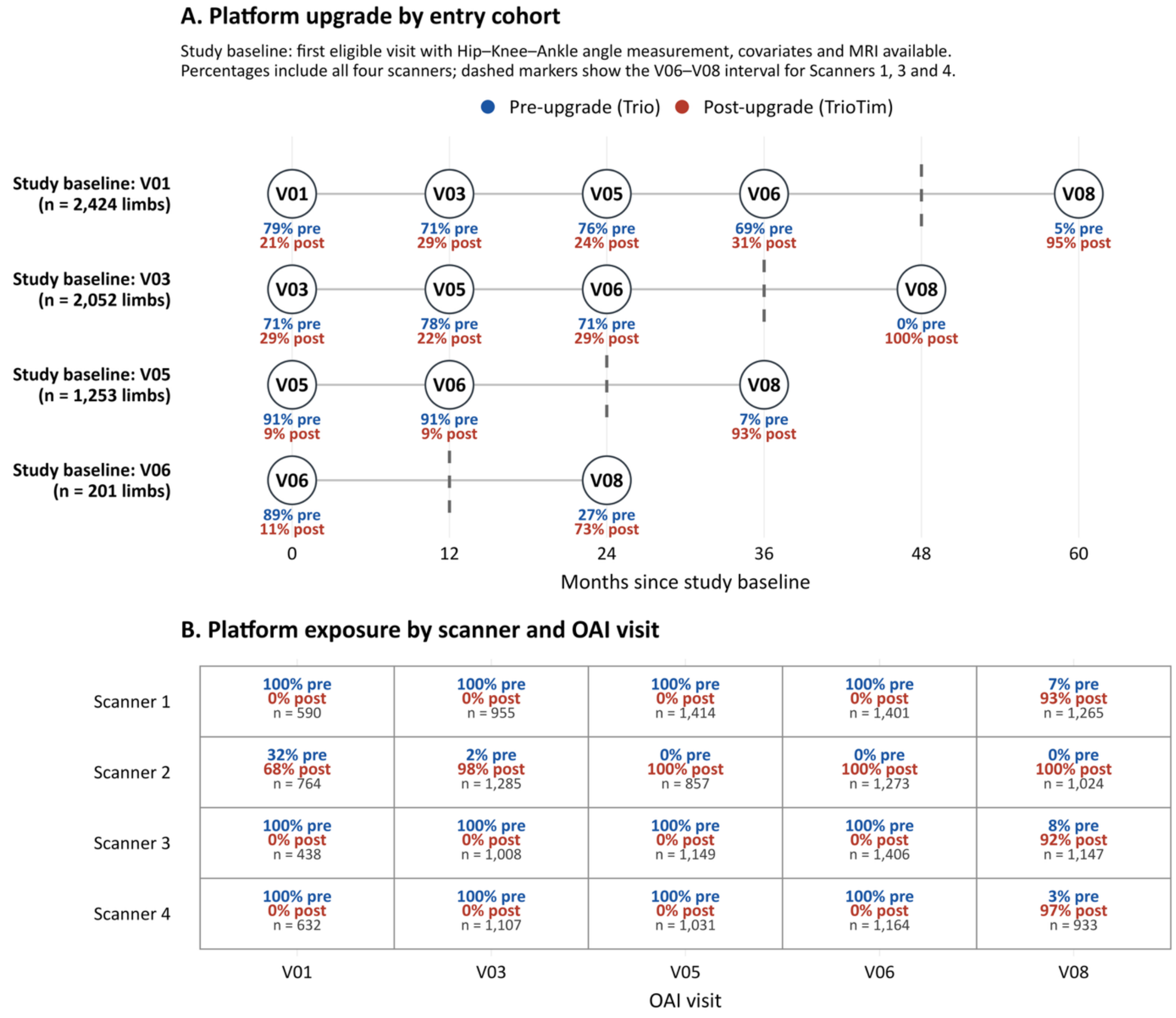


**Supplementary Figure 1. Scanner-platform timing in the analytic cohort.** Both panels show MRI examinations from the current study's analytic cohort, at or after each limb's study baseline. Blue denotes pre-upgrade examinations (Trio), and red denotes post-upgrade examinations (TrioTim). Panel A groups limbs by their study baseline, defined as the first eligible visit with Hip-Knee-Ankle angle measurement, covariates and MRI available, and plots follow-up time from that visit. Percentages summarize the available examinations across all scanners at each visit. Dashed gray markers indicate the V06-V08 upgrade interval for Scanners 1, 3 and 4. Scanner 2 upgraded earlier than the other three scanners, as shown by the predominantly post-upgrade examinations at V01 and V03 in panel B. Panel B shows platform percentages and MRI examination counts separately for each scanner and OAI visit.

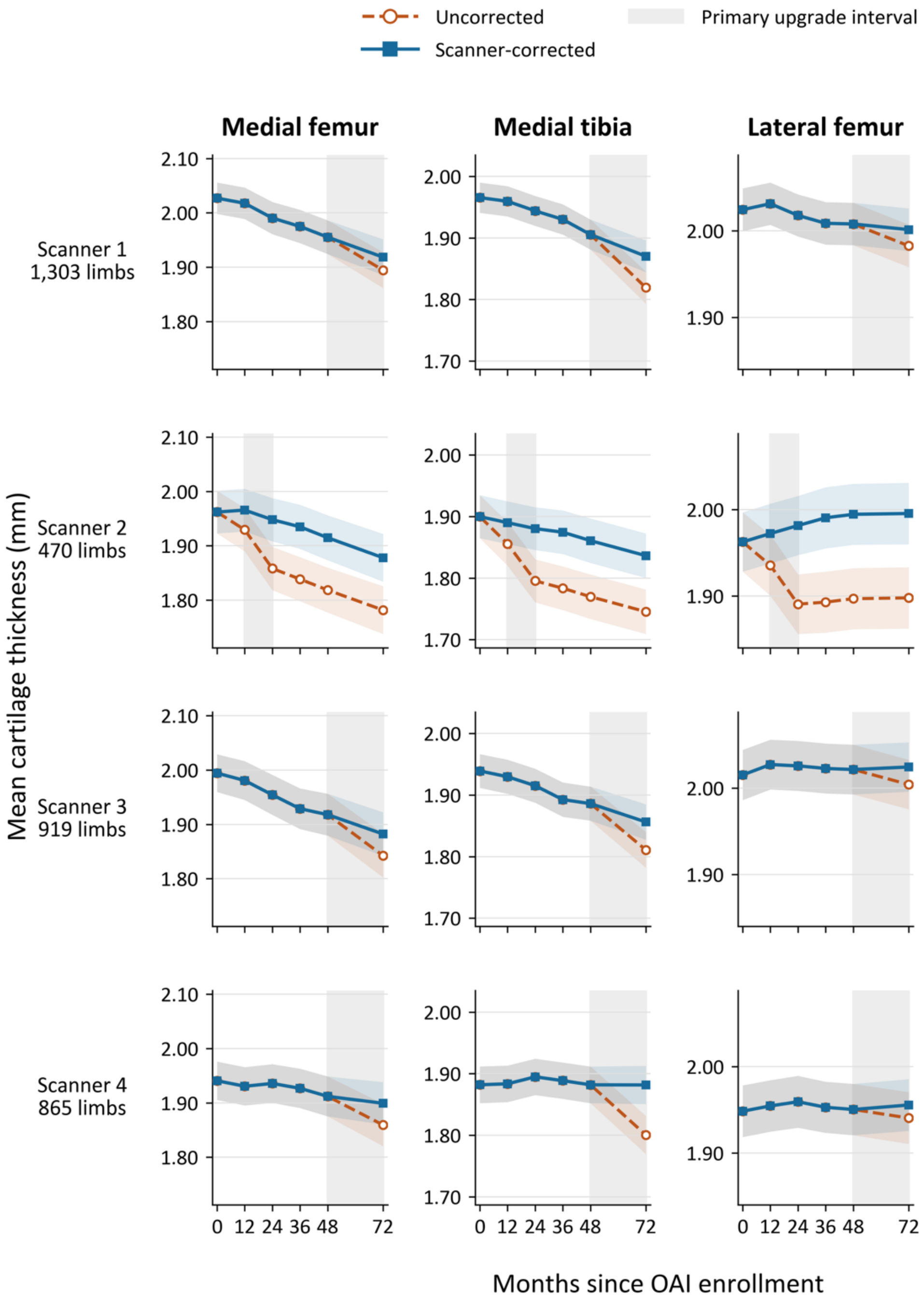


**Supplementary Figure 2. Cartilage Thickness Before and After Scanner Correction.** Mean cartilage thickness is shown across OAI visits for 3,557 limbs with measurements in all four Regions at every displayed visit. Orange dashed lines show uncorrected cartilage measurements, and blue solid lines show cartilage measurements after correction. Colored bands indicate pointwise 95% confidence intervals clustered by participant, treating the correction offsets as fixed. Grey shading marks the primary upgrade interval for each Scanner. Correction reduced the upgrade-related decreases in measured cartilage thickness.

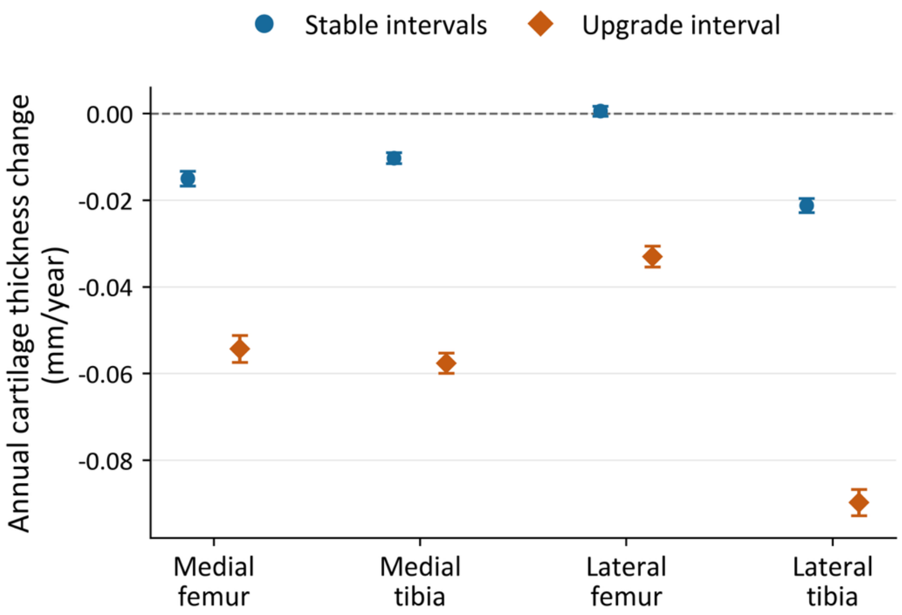


**Supplementary Figure 3. Annual Cartilage Thickness Change During Stable and Upgrade Intervals.** Points show mean uncorrected annual cartilage thickness change and participant-clustered 95% confidence intervals, pooled across scanners in the broader OAI imaging cohort after quality exclusions. Blue circles represent intervals with an unchanged scanner platform, and orange diamonds represent intervals spanning the Trio-to-TrioTim upgrade. Rates were calculated between adjacent scheduled visits at 0, 12, 24, 36, 48, and 72 months, dividing thickness change by interval duration in years. Each region included 4,944 to 4,946 limbs with both interval types. Stable rates were averaged within each limb before calculating equally limb-weighted means. Negative values indicate thinning. Greater apparent thinning during upgrade intervals in all four regions supports accounting for the scanner upgrade. To estimate correction offsets, we calculated each eligible limb's expected thickness change during its upgrade interval by multiplying its mean stable-interval annual rate by the upgrade-interval duration. The limb-specific difference was the observed upgrade-interval change minus this expected change. We then averaged these differences within each scanner and region, producing 16 correction offsets. All 16 offsets were negative, ranging from −0.1191 to −0.0157 mm, and all were statistically significant (Supplementary Table 2). We retained all four scanners and subtracted the applicable offset from each post-upgrade cartilage measurement before model fitting.

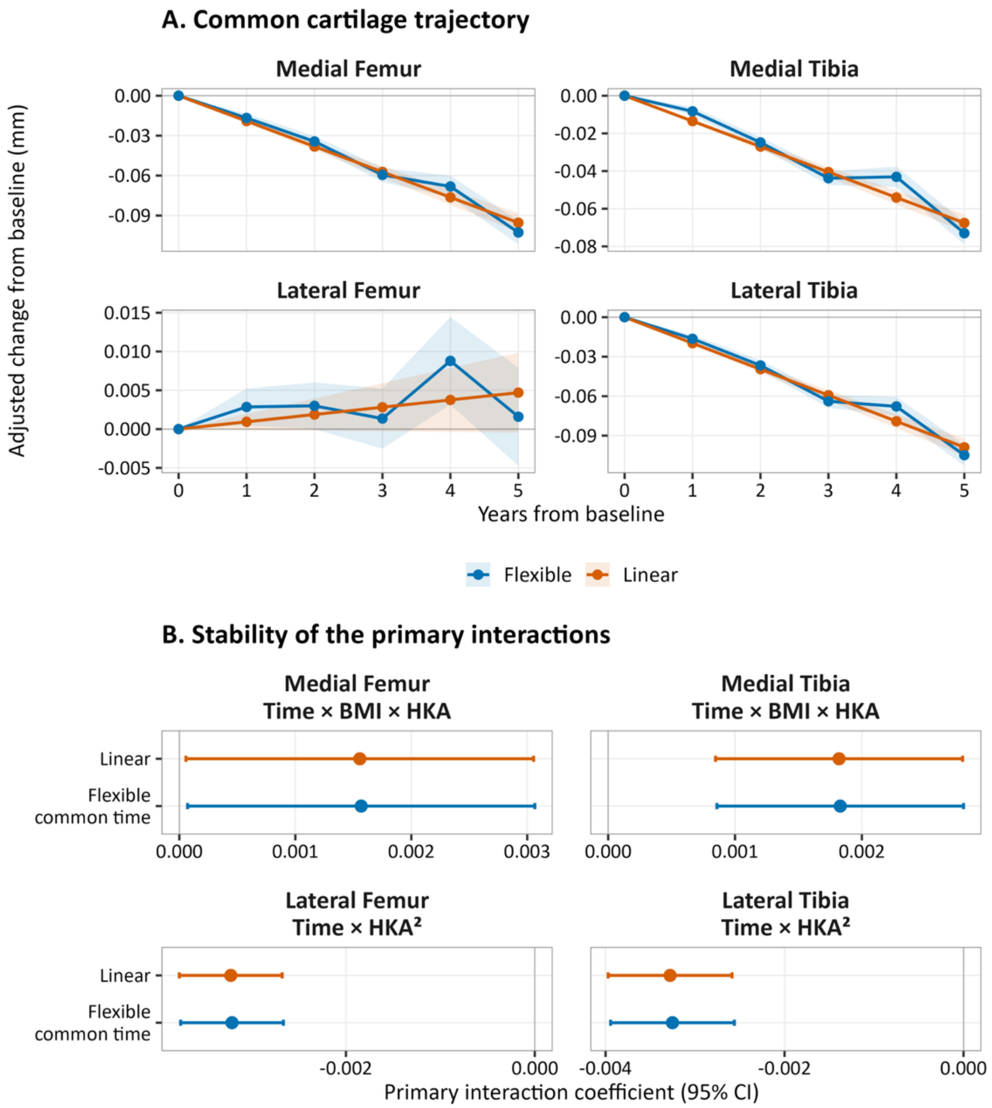


**Supplementary Figure 4A. Evaluation of the Linearity Assumptions for Time.** Comparison of continuous linear time with a flexible common time trajectory in the corrected primary models. The upper panels compare the estimated common cartilage trajectories. The lower panels compare estimates and 95% confidence intervals for the primary interactions: time × BMI × HKA in the medial femur and medial tibia, and time × HKA² in the lateral femur and lateral tibia. Allowing the common trajectory to vary across visits changed each primary interaction estimate by less than 1%, supporting continuous linear time for estimating average annual differences in cartilage thickness change.

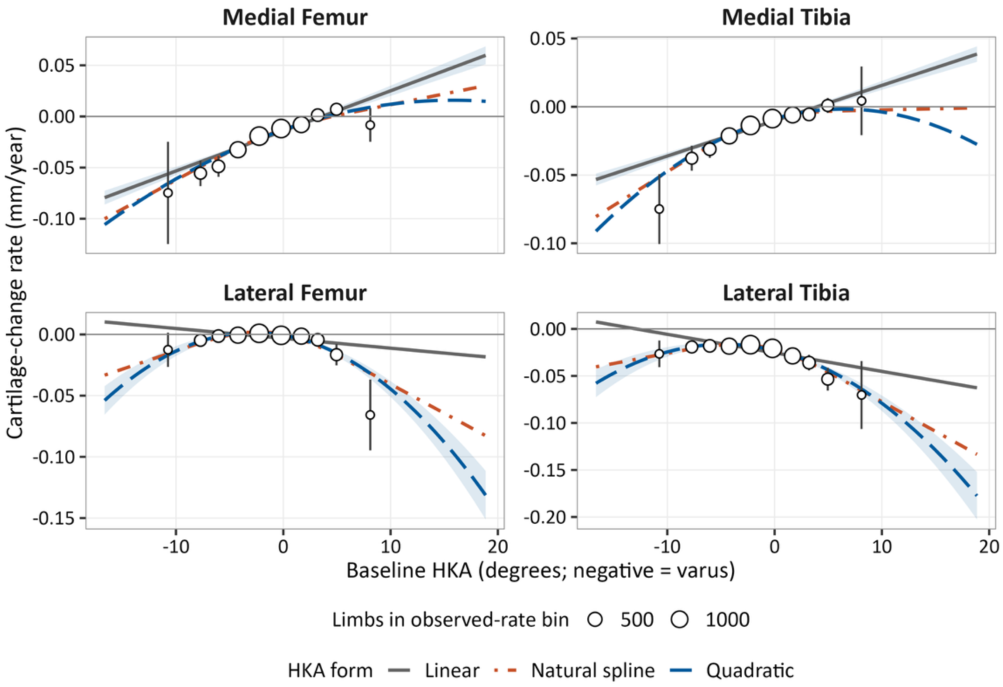


**Supplementary Figure 4B. Evaluation of the Linearity Assumptions for Alignment.** Rates calculated separately for each limb are compared with rates estimated using linear, quadratic, and natural-spline HKA terms in the corrected models. Points show limb-level rates summarized across populated HKA bins, with point size indicating the number of limbs and error bars indicating 95% confidence intervals. HKA models with nonlinear terms more closely matched the observed rates, particularly in the medial tibia and lateral regions. The medial relationships generally showed less thinning from varus toward valgus, with some flattening in the medial tibia. Linear HKA was retained to summarize the average association between alignment and medial thinning and its interaction with BMI. A separate medial-tibial sensitivity model added only $HKA^2$ and time × $HKA^2$ to the primary model. The time × BMI × HKA interaction remained positive and significant ($\beta = 0.0015$, 95% CI [0.0005, 0.0025], $p = 0.003$). The comparison of predicted five-year changes and the rationale for retaining linear medial HKA are reported in Supplementary Table 8. In the lateral regions, quadratic HKA (time × $HKA^2$) represented faster thinning toward both alignment extremes.

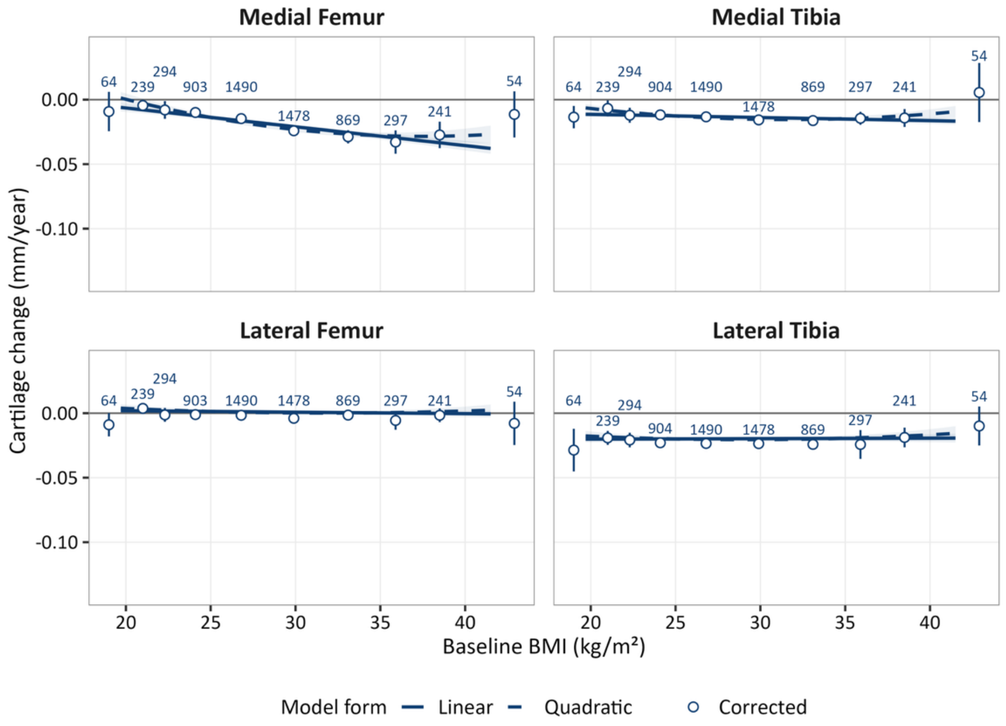


**Supplementary Figure 4C. Evaluation of the Linearity Assumptions for BMI.** Rates calculated separately for each limb are compared with rates estimated using linear and quadratic BMI terms in the corrected models. Points show limb-level rates summarized across populated BMI bins, labels indicate the number of limbs, and error bars indicate 95% confidence intervals.

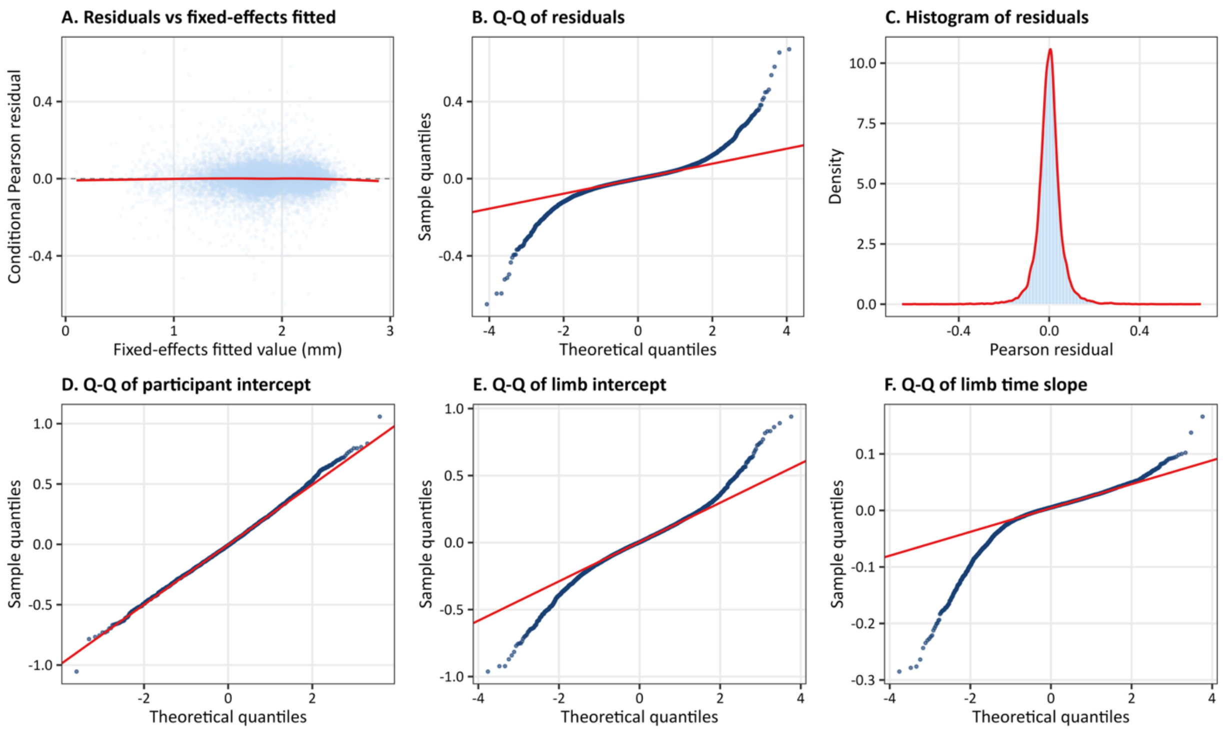


**Supplementary Figure 5A. Primary Model Diagnostics for the Medial Femur.** Diagnostics are shown for the corrected primary linear-HKA model. Panels show (A) conditional Pearson residuals versus cartilage thickness predicted from the fixed effects only, with a locally fitted trend; (B) a quantile-quantile plot of the Pearson residuals; (C) the residual distribution with a density curve; and quantile-quantile plots of the (D) participant intercept, (E) limb intercept, and (F) limb-specific time slope. The residuals remained centered near zero across most fitted values, with modest heteroscedasticity and heavier-than-normal tails in the residual and random-effect distributions.

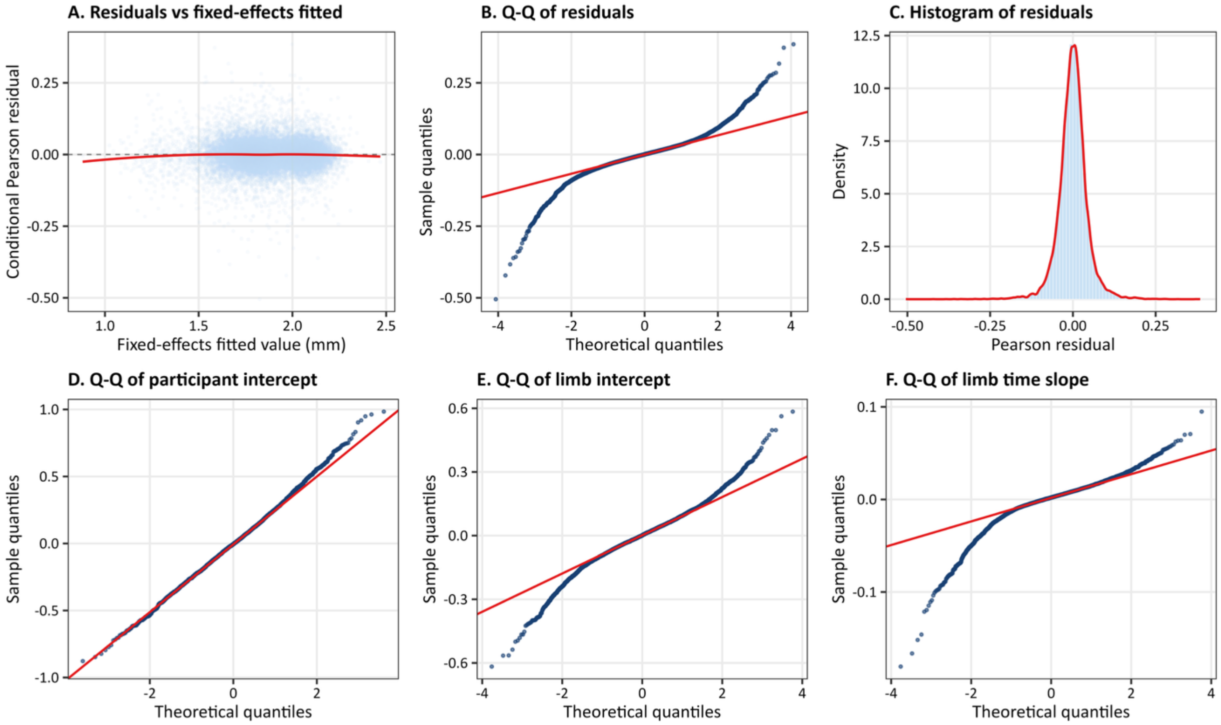


**Supplementary Figure 5B. Primary Model Diagnostics for the Medial Tibia.** Diagnostics are shown for the corrected primary linear-HKA model. Panels show (A) conditional Pearson residuals versus cartilage thickness predicted from the fixed effects only, with a locally fitted trend; (B) a quantile-quantile plot of the Pearson residuals; (C) the residual distribution with a density curve; and quantile-quantile plots of the (D) participant intercept, (E) limb intercept, and (F) limb-specific time slope. The residuals remained centered near zero across most fitted values, with modest departures from constant residual variance and normality, primarily in the distribution tails.

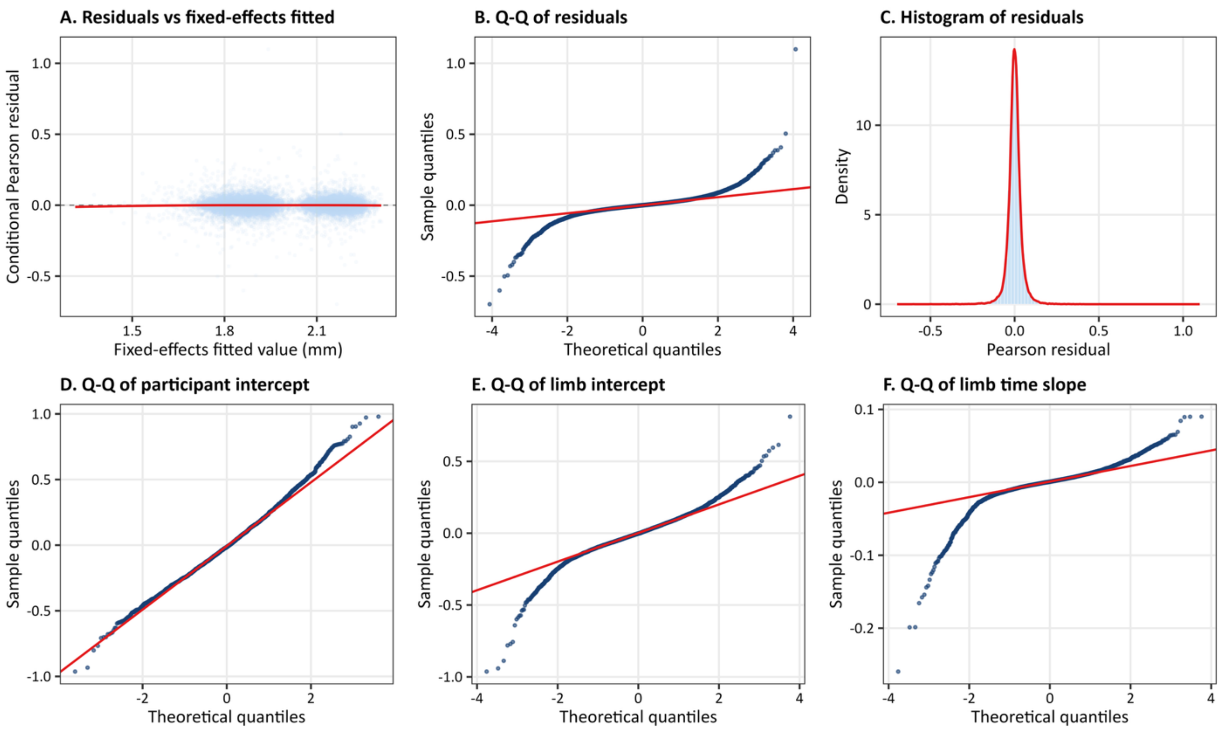


**Supplementary Figure 5C. Primary Model Diagnostics for the Lateral Femur.** Diagnostics are shown for the corrected primary quadratic-HKA model. Panels show (A) conditional Pearson residuals versus cartilage thickness predicted from the fixed effects only, with a locally fitted trend; (B) a quantile-quantile plot of the Pearson residuals; (C) the residual distribution with a density curve; and quantile-quantile plots of the (D) participant intercept, (E) limb intercept, and (F) limb-specific time slope. The residuals remained centered near zero across most fitted values, although the residual and random-effect distributions showed heavier-than-normal tails.

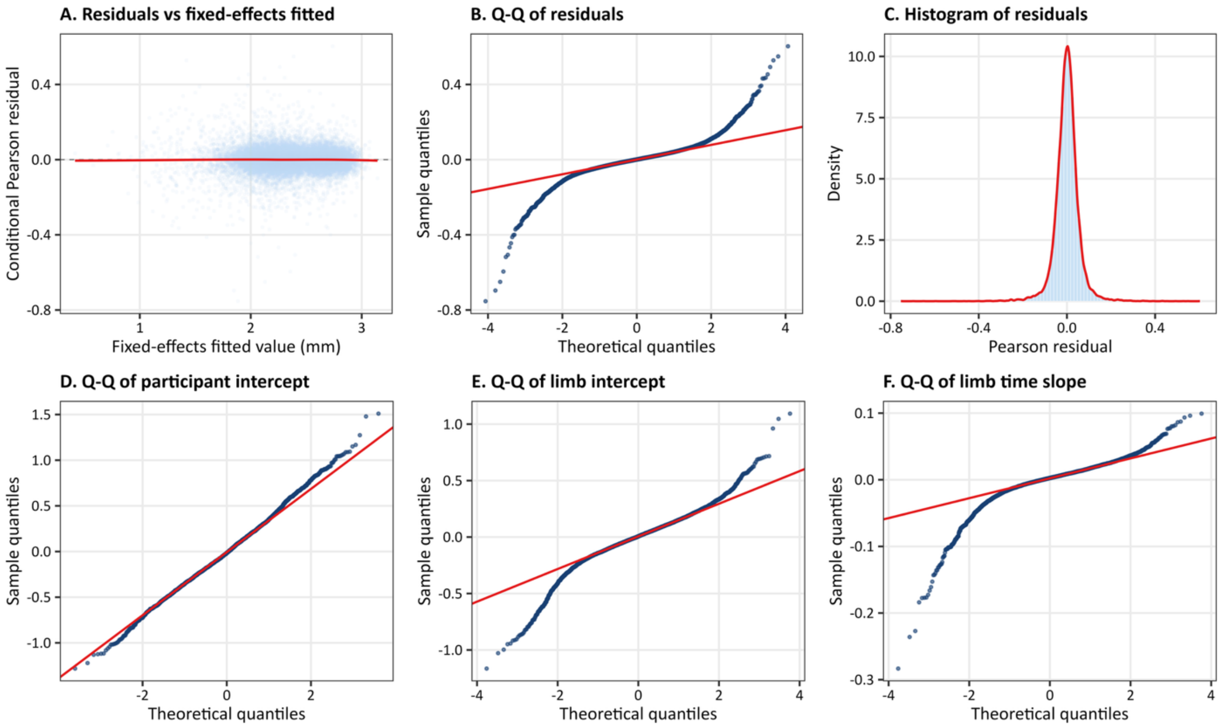


**Supplementary Figure 5D. Primary Model Diagnostics for the Lateral Tibia.** Diagnostics are shown for the corrected primary quadratic-HKA model. Panels show (A) conditional Pearson residuals versus cartilage thickness predicted from the fixed effects only, with a locally fitted trend; (B) a quantile-quantile plot of the Pearson residuals; (C) the residual distribution with a density curve; and quantile-quantile plots of the (D) participant intercept, (E) limb intercept, and (F) limb-specific time slope. The residuals remained centered near zero across most fitted values, with some nonconstant residual variance and heavier-than-normal tails in the residual and random-effect distributions.

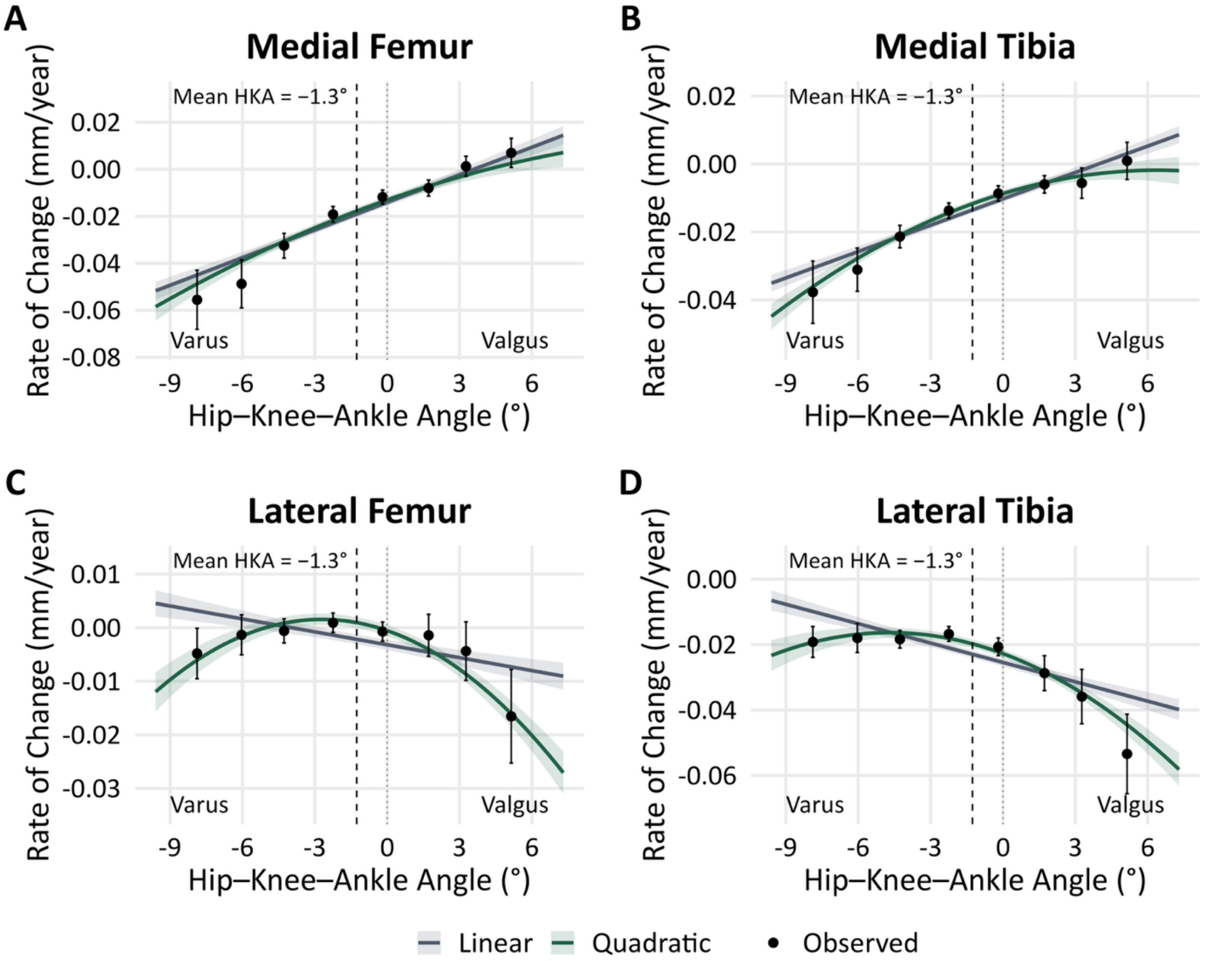


**Supplementary Figure 6. Observed and Model-Predicted Cartilage Thickness Change Across Alignment.** Annual cartilage thickness change is shown for the medial femur (A), medial tibia (B), lateral femur (C), and lateral tibia (D). For the observed data, limbs were grouped by baseline HKA into percentile ranges: 1st–5th, 5th–10th, 10th–25th, 25th–50th, 50th–75th, 75th–90th, 90th–95th, and 95th–99th. Each black point shows the group's mean HKA on the horizontal axis and mean observed annual cartilage thickness change on the vertical axis. Annual change was estimated separately for each limb from its available thickness measurements over time. Error bars represent 95% confidence intervals for the group means using participant-clustered standard errors. Grey and green lines show adjusted linear and quadratic HKA model predictions at mean BMI, respectively; shading represents pointwise 95% confidence intervals. Observed points are unadjusted summaries. The dashed vertical line marks mean HKA (−1.3°), and the dotted line marks 0°. Negative rates indicate thinning. The quadratic curves illustrate faster lateral thinning toward both alignment extremes. Primary analyses retain linear HKA medially and quadratic HKA laterally.

| Region | Paired readings | Pearson r | Average Automated (mm) | Average Manual (mm) | Difference (mm) | Automated SRM | Manual SRM |
|---|---|---|---|---|---|---|---|
| Medial femur | 1,199 | 0.925 | 1.84 | 1.66 | 0.17 | -0.578 | -0.455 |
| Lateral femur | 1,199 | 0.909 | 2.04 | 1.93 | 0.11 | -0.064 | 0.081 |
| Medial tibia | 1,199 | 0.915 | 1.90 | 1.61 | 0.29 | -0.418 | -0.371 |
| Lateral tibia | 1,199 | 0.943 | 2.43 | 1.94 | 0.49 | -0.546 | -0.393 |

**Supplementary Table 1. Agreement and longitudinal sensitivity of automated and manual cartilage thickness measurements.** Automated (deep-learning segmentation) and manual weight-bearing cartilage thickness were compared in 1,199 knee-visits from 600 knees of the Foundation for the National Institutes of Health (FNIH) OAI sub-cohort, at the baseline and 24-month visits (the timepoints with paired manual readings). Pearson r reflects cross-sectional correlation; the standardized response mean (SRM) reflects sensitivity to longitudinal change (baseline to 24 months), with larger magnitude indicating greater sensitivity. Cross-sectional correlations ranged from 0.91 to 0.94. Automated measurements showed greater responsiveness to longitudinal change than manual measurements in 3 of 4 Regions.

| Region | Scanner | Estimation limbs | Estimated offset (mm) | 95% CI (mm) |
|---|---|---|---|---|
| Medial femur | Scanner 1 | 1,388 | -0.0260 | [-0.0344, -0.0176] |
| | Scanner 2 | 1,340 | -0.0963 | [-0.1050, -0.0877] |
| | Scanner 3 | 1,177 | -0.0438 | [-0.0543, -0.0333] |
| | Scanner 4 | 1,041 | -0.0416 | [-0.0522, -0.0310] |
| Medial tibia | Scanner 1 | 1,388 | -0.0554 | [-0.0620, -0.0489] |
| | Scanner 2 | 1,340 | -0.0911 | [-0.0985, -0.0836] |
| | Scanner 3 | 1,177 | -0.0501 | [-0.0580, -0.0423] |
| | Scanner 4 | 1,041 | -0.0847 | [-0.0920, -0.0774] |
| Lateral femur | Scanner 1 | 1,386 | -0.0203 | [-0.0265, -0.0142] |
| | Scanner 2 | 1,340 | -0.0975 | [-0.1043, -0.0907] |
| | Scanner 3 | 1,177 | -0.0223 | [-0.0286, -0.0160] |
| | Scanner 4 | 1,041 | -0.0157 | [-0.0227, -0.0087] |
| Lateral tibia | Scanner 1 | 1,388 | -0.1053 | [-0.1130, -0.0976] |
| | Scanner 2 | 1,340 | -0.1191 | [-0.1298, -0.1085] |
| | Scanner 3 | 1,177 | -0.0915 | [-0.0997, -0.0833] |
| | Scanner 4 | 1,041 | -0.0984 | [-0.1078, -0.0889] |

**Supplementary Table 2. Scanner platform measurement offsets.** Offsets were estimated separately for each scanner and cartilage region using limbs with an observed upgrade interval and at least one stable interval. Annualized changes across all eligible adjacent scheduled stable intervals through V08, including post-upgrade intervals, were averaged within each limb. The limb offset was the observed upgrade-interval change minus this mean stable rate multiplied by the upgrade-interval duration. Limb offsets were then averaged within each scanner and region. Negative values indicate lower measured cartilage thickness after the upgrade before correction. The applicable offset was subtracted from each post-upgrade measurement.

| Medial Femur | | | |
|---|---|---|---|
| **Term** | **β** | **95% CI** | **P Value** |
| Intercept | +2.2119 | [+2.1902, +2.2335] | <0.001 |
| **Main effects** | | | |
| Time (years) | -0.0191 | [-0.0205, -0.0177] | <0.001 |
| BMI (per SD) | -0.0013 | [-0.0134, +0.0108] | 0.834 |
| HKA angle (per SD) | +0.1491 | [+0.1393, +0.1588] | <0.001 |
| Age (per SD) | -0.0344 | [-0.0463, -0.0225] | <0.001 |
| Sex (female vs male) | -0.4468 | [-0.4707, -0.4228] | <0.001 |
| **KL grade** | | | |
| KL 1 (vs KL 0) | +0.0231 | [-0.0012, +0.0475] | 0.063 |
| KL 2 (vs KL 0) | +0.0221 | [-0.0005, +0.0448] | 0.055 |
| KL 3 (vs KL 0) | -0.2646 | [-0.2915, -0.2377] | <0.001 |
| KL 4 (vs KL 0) | -0.7620 | [-0.8017, -0.7222] | <0.001 |
| **Interactions** | | | |
| Time x BMI | -0.0070 | [-0.0084, -0.0055] | <0.001 |
| Time x HKA angle | +0.0132 | [+0.0117, +0.0146] | <0.001 |
| BMI x HKA angle | +0.0209 | [+0.0113, +0.0306] | <0.001 |
| Time x BMI x HKA angle | +0.0016 | [+0.0001, +0.0031] | 0.042 |
| **Medial Tibia** | | | |
| **Term** | **β** | **95% CI** | **P Value** |
| Intercept | +2.0384 | [+2.0201, +2.0567] | <0.001 |
| **Main effects** | | | |
| Time (years) | -0.0135 | [-0.0144, -0.0126] | <0.001 |
| BMI (per SD) | +0.0177 | [+0.0069, +0.0284] | 0.001 |
| HKA angle (per SD) | +0.0762 | [+0.0690, +0.0835] | <0.001 |
| Age (per SD) | -0.0421 | [-0.0528, -0.0315] | <0.001 |
| Sex (female vs male) | -0.2975 | [-0.3189, -0.2761] | <0.001 |
| **KL grade** | | | |
| KL 1 (vs KL 0) | +0.0249 | [+0.0075, +0.0423] | 0.005 |
| KL 2 (vs KL 0) | +0.0513 | [+0.0347, +0.0678] | <0.001 |
| KL 3 (vs KL 0) | -0.0475 | [-0.0671, -0.0279] | <0.001 |
| KL 4 (vs KL 0) | -0.3452 | [-0.3733, -0.3171] | <0.001 |
| **Interactions** | | | |
| Time x BMI | -0.0012 | [-0.0021, -0.0002] | 0.014 |
| Time x HKA angle | +0.0087 | [+0.0078, +0.0096] | <0.001 |
| BMI x HKA angle | +0.0121 | [+0.0049, +0.0193] | <0.001 |
| Time x BMI x HKA angle | +0.0018 | [+0.0008, +0.0028] | <0.001 |

**Supplementary Table 3A. Primary Linear Mixed-Effects Model Results for Medial Femur and Medial Tibia Cartilage Thickness.** Coefficients are in mm for terms without time and mm/year for terms involving time. BMI, HKA, and age are expressed in SD units. The time x BMI x HKA interaction was positive in the medial femur (β = 0.0016 mm/year per 1-SD BMI * 1-SD HKA, 95% CI [0.0001, 0.0031], p = 0.042) and medial tibia (β = 0.0018, 95% CI [0.0008, 0.0028], p < 0.001), indicating that BMI and HKA alignment interact to modify the longitudinal rate of cartilage loss in the medial region.

| Lateral Femur | | | |
|---|---|---|---|
| **Term** | **β** | **95% CI** | **P Value** |
| Intercept | +2.1311 | [+2.1125, +2.1498] | <0.001 |
| **Main effects** | | | |
| Time (years) | +0.0009 | [-0.0001, +0.0020] | 0.074 |
| BMI (per SD) | +0.0279 | [+0.0173, +0.0386] | <0.001 |
| HKA angle (per SD) | -0.0229 | [-0.0306, -0.0153] | <0.001 |
| $HKA^2$ (per $SD^2$) | -0.0093 | [-0.0132, -0.0054] | <0.001 |
| Age (per SD) | -0.0147 | [-0.0253, -0.0042] | 0.006 |
| Sex (female vs male) | -0.3002 | [-0.3215, -0.2790] | <0.001 |
| **KL grade** | | | |
| KL 1 (vs KL 0) | +0.0298 | [+0.0112, +0.0485] | 0.002 |
| KL 2 (vs KL 0) | +0.0798 | [+0.0622, +0.0974] | <0.001 |
| KL 3 (vs KL 0) | +0.0487 | [+0.0274, +0.0700] | <0.001 |
| KL 4 (vs KL 0) | -0.0294 | [-0.0624, +0.0036] | 0.081 |
| **Interactions** | | | |
| Time x BMI | -0.0005 | [-0.0014, +0.0004] | 0.242 |
| Time x HKA angle | -0.0028 | [-0.0037, -0.0019] | <0.001 |
| Time x $HKA^2$ | -0.0032 | [-0.0038, -0.0027] | <0.001 |
| BMI x HKA angle | +0.0109 | [+0.0034, +0.0185] | 0.005 |
| **Lateral Tibia** | | | |
| **Term** | **β** | **95% CI** | **P Value** |
| Intercept | +2.6740 | [+2.6469, +2.7010] | <0.001 |
| **Main effects** | | | |
| Time (years) | -0.0198 | [-0.0211, -0.0185] | <0.001 |
| BMI (per SD) | +0.0223 | [+0.0067, +0.0378] | 0.005 |
| HKA angle (per SD) | -0.1345 | [-0.1455, -0.1234] | <0.001 |
| $HKA^2$ (per $SD^2$) | -0.0131 | [-0.0187, -0.0074] | <0.001 |
| Age (per SD) | -0.0807 | [-0.0960, -0.0653] | <0.001 |
| Sex (female vs male) | -0.3840 | [-0.4149, -0.3531] | <0.001 |
| **KL grade** | | | |
| KL 1 (vs KL 0) | -0.0284 | [-0.0550, -0.0018] | 0.037 |
| KL 2 (vs KL 0) | -0.0702 | [-0.0955, -0.0450] | <0.001 |
| KL 3 (vs KL 0) | -0.2764 | [-0.3068, -0.2460] | <0.001 |
| KL 4 (vs KL 0) | -0.5977 | [-0.6448, -0.5506] | <0.001 |
| **Interactions** | | | |
| Time x BMI | +0.0002 | [-0.0010, +0.0013] | 0.757 |
| Time x HKA angle | -0.0067 | [-0.0078, -0.0056] | <0.001 |
| Time x $HKA^2$ | -0.0033 | [-0.0040, -0.0026] | <0.001 |
| BMI x HKA angle | +0.0051 | [-0.0057, +0.0159] | 0.353 |

**Supplementary Table 3B. Primary Linear Mixed-Effects Model Results for Lateral Femur and Lateral Tibia Cartilage Thickness.** Coefficients involving BMI or linear HKA are reported per 1-SD difference in the corresponding variable; the time x $HKA^2$ coefficient is reported per squared standardized HKA unit. The time x $HKA^2$ interaction was negative in the lateral femur ($\beta = -0.0032$ mm/year, 95% CI [−0.0038, −0.0027], $p < 0.001$) and lateral tibia ($\beta = -0.0033$, 95% CI [−0.0040, −0.0026], $p < 0.001$), whereas the time x BMI interactions were not statistically significant. These findings indicate a nonlinear association between HKA alignment and lateral cartilage loss, without evidence that BMI modified the lateral rate of loss.

| Medial Femur | | | |
|---|---|---|---|
| **Term** | **β** | **95% CI** | **p** |
| Intercept | +2.1849 | [+2.1630, +2.2067] | <0.001 |
| Main effects | | | |
| Time (years) | -0.0257 | [-0.0271, -0.0243] | <0.001 |
| BMI (per SD) | -0.0024 | [-0.0147, +0.0098] | 0.698 |
| HKA angle (per SD) | +0.1491 | [+0.1393, +0.1589] | <0.001 |
| Age (per SD) | -0.0321 | [-0.0441, -0.0201] | <0.001 |
| Sex (female vs male) | -0.4502 | [-0.4745, -0.4260] | <0.001 |
| **KL grade** | | | |
| KL 1 (vs KL 0) | +0.0225 | [-0.0020, +0.0469] | 0.072 |
| KL 2 (vs KL 0) | +0.0197 | [-0.0030, +0.0425] | 0.089 |
| KL 3 (vs KL 0) | -0.2657 | [-0.2928, -0.2387] | <0.001 |
| KL 4 (vs KL 0) | -0.7621 | [-0.8020, -0.7222] | <0.001 |
| **Interactions** | | | |
| Time x BMI | -0.0067 | [-0.0082, -0.0053] | <0.001 |
| Time x HKA angle | +0.0131 | [+0.0117, +0.0146] | <0.001 |
| BMI x HKA angle | +0.0213 | [+0.0116, +0.0310] | <0.001 |
| Time x BMI x HKA angle | +0.0015 | [-0.0000, +0.0030] | 0.057 |
| **Medial Tibia** | | | |
| Term | β | 95% CI | p |
| Intercept | +2.0084 | [+1.9898, +2.0270] | <0.001 |
| **Main effects** | | | |
| Time (years) | -0.0241 | [-0.0250, -0.0232] | <0.001 |
| BMI (per SD) | +0.0169 | [+0.0060, +0.0279] | 0.002 |
| HKA angle (per SD) | +0.0761 | [+0.0688, +0.0833] | <0.001 |
| Age (per SD) | -0.0403 | [-0.0511, -0.0294] | <0.001 |
| Sex (female vs male) | -0.3008 | [-0.3227, -0.2790] | <0.001 |
| **KL grade** | | | |
| KL 1 (vs KL 0) | +0.0244 | [+0.0069, +0.0419] | 0.006 |
| KL 2 (vs KL 0) | +0.0495 | [+0.0328, +0.0661] | <0.001 |
| KL 3 (vs KL 0) | -0.0499 | [-0.0696, -0.0302] | <0.001 |
| KL 4 (vs KL 0) | -0.3473 | [-0.3755, -0.3190] | <0.001 |
| **Interactions** | | | |
| Time x BMI | -0.0007 | [-0.0016, +0.0003] | 0.158 |
| Time x HKA angle | +0.0086 | [+0.0077, +0.0096] | <0.001 |
| BMI x HKA angle | +0.0123 | [+0.0051, +0.0196] | <0.001 |
| Time x BMI x HKA angle | +0.0017 | [+0.0008, +0.0027] | <0.001 |

**Supplementary Table 4A. Uncorrected Linear Mixed-Effects Model Results for Medial Femur and Medial Tibia Cartilage Thickness.** Interaction coefficients involving BMI or HKA are reported per 1-SD difference in the corresponding variable. The time x BMI x HKA interaction was positive in the medial femur (β = 0.0015 mm/year per 1-SD BMI * 1-SD HKA, 95% CI [−0.0000, 0.0030], p = 0.057) and medial tibia (β = 0.0017, 95% CI [0.0008, 0.0027], p < 0.001). Compared with the corrected models, these estimates were 6% and 4% smaller, respectively. The medial tibial interaction remained statistically significant, whereas the medial femoral interaction changed from p = 0.042 to p = 0.057. Thus, the direction and magnitude of the medial interactions were similar without scanner correction, although the medial femur interaction was no longer significant.

| Lateral Femur | | | |
|---|---|---|---|
| **Term** | **β** | **95% CI** | **p** |
| Intercept | +2.1065 | [+2.0876, +2.1255] | <0.001 |
| **Main effects** | | | |
| Time (years) | -0.0030 | [-0.0041, -0.0020] | <0.001 |
| BMI (per SD) | +0.0265 | [+0.0157, +0.0374] | <0.001 |
| HKA angle (per SD) | -0.0241 | [-0.0318, -0.0164] | <0.001 |
| $HKA^2$ (per $SD^2$) | -0.0095 | [-0.0134, -0.0056] | <0.001 |
| Age (per SD) | -0.0122 | [-0.0230, -0.0014] | 0.027 |
| Sex (female vs male) | -0.3026 | [-0.3243, -0.2809] | <0.001 |
| **KL grade** | | | |
| KL 1 (vs KL 0) | +0.0289 | [+0.0102, +0.0476] | 0.002 |
| KL 2 (vs KL 0) | +0.0772 | [+0.0595, +0.0949] | <0.001 |
| KL 3 (vs KL 0) | +0.0464 | [+0.0250, +0.0678] | <0.001 |
| KL 4 (vs KL 0) | -0.0320 | [-0.0651, +0.0012] | 0.059 |
| **Interactions** | | | |
| Time x BMI | -0.0005 | [-0.0014, +0.0004] | 0.300 |
| Time x HKA angle | -0.0028 | [-0.0037, -0.0018] | <0.001 |
| Time x $HKA^2$ | -0.0033 | [-0.0038, -0.0027] | <0.001 |
| BMI x HKA angle | +0.0111 | [+0.0035, +0.0188] | 0.004 |
| **Lateral Tibia** | | | |
| **Term** | **β** | **95% CI** | **p** |
| Intercept | +2.6309 | [+2.6036, +2.6583] | <0.001 |
| **Main effects** | | | |
| Time (years) | -0.0368 | [-0.0381, -0.0354] | <0.001 |
| BMI (per SD) | +0.0218 | [+0.0061, +0.0376] | 0.007 |
| HKA angle (per SD) | -0.1356 | [-0.1467, -0.1245] | <0.001 |
| $HKA^2$ (per $SD^2$) | -0.0124 | [-0.0180, -0.0068] | <0.001 |
| Age (per SD) | -0.0779 | [-0.0934, -0.0623] | <0.001 |
| Sex (female vs male) | -0.3884 | [-0.4197, -0.3570] | <0.001 |
| **KL grade** | | | |
| KL 1 (vs KL 0) | -0.0293 | [-0.0560, -0.0026] | 0.032 |
| KL 2 (vs KL 0) | -0.0729 | [-0.0982, -0.0475] | <0.001 |
| KL 3 (vs KL 0) | -0.2786 | [-0.3092, -0.2481] | <0.001 |
| KL 4 (vs KL 0) | -0.6013 | [-0.6486, -0.5541] | <0.001 |
| **Interactions** | | | |
| Time x BMI | +0.0012 | [+0.0000, +0.0024] | 0.050 |
| Time x HKA angle | -0.0068 | [-0.0080, -0.0056] | <0.001 |
| Time x $HKA^2$ | -0.0029 | [-0.0036, -0.0021] | <0.001 |
| BMI x HKA angle | +0.0054 | [-0.0055, +0.0163] | 0.333 |

**Supplementary Table 4B. Uncorrected Linear Mixed-Effects Model Results for Lateral Femur and Lateral Tibia Cartilage Thickness.** Coefficients involving BMI or linear HKA are reported per 1-SD difference in the corresponding variable; the time x $HKA^2$ coefficient is reported per squared standardized HKA unit. The time x $HKA^2$ interaction was negative in the lateral femur ($\beta = -0.0033$ mm/year, 95% CI [−0.0038, −0.0027], $p < 0.001$) and lateral tibia ($\beta = -0.0029$, 95% CI [−0.0036, −0.0021], $p < 0.001$). Compared with the corrected models, these estimates differed by 2% and 13%, respectively, and both remained statistically significant, supporting a nonlinear association between HKA alignment and lateral cartilage loss. The lateral femoral time x BMI interaction remained non-significant ($p = 0.300$), whereas the small lateral tibial time x BMI estimate increased from $\beta = 0.0002$ ($p = 0.757$) in the corrected model to $\beta = 0.0012$ ($p = 0.050$) without correction. The lateral $HKA^2$ findings were robust, while the lateral tibial time x BMI estimate was more sensitive to scanner correction.

| HKA Percentile<br>Angle (°) | BMI 1st<br>20 kg/m² | BMI 25th<br>25 kg/m² | BMI 50th<br>28 kg/m² | BMI 75th<br>32 kg/m² | BMI 99th<br>42 kg/m² |
|---|---|---|---|---|---|
| **1st<br>-9.6** | 0y<br>1.99 [1.94, 2.05]<br>5y<br>1.83 [1.77, 1.90] | 0y<br>1.96 [1.92, 1.99]<br>5y<br>1.74 [1.70, 1.78] | 0y<br>1.94 [1.90, 1.97]<br>5y<br>1.68 [1.65, 1.72] | 0y<br>1.91 [1.87, 1.94]<br>5y<br>1.61 [1.57, 1.65] | 0y<br>1.83 [1.75, 1.91]<br>5y<br>1.42 [1.33, 1.52] |
| **25th<br>-3.4** | 0y<br>2.17 [2.13, 2.20]<br>5y<br>2.10 [2.07, 2.14] | 0y<br>2.16 [2.14, 2.19]<br>5y<br>2.06 [2.03, 2.08] | 0y<br>2.16 [2.14, 2.19]<br>5y<br>2.03 [2.01, 2.06] | 0y<br>2.16 [2.14, 2.19]<br>5y<br>2.00 [1.97, 2.02] | 0y<br>2.16 [2.12, 2.21]<br>5y<br>1.91 [1.86, 1.96] |
| **50th<br>-1.2** | 0y<br>2.23 [2.20, 2.26]<br>5y<br>2.20 [2.16, 2.23] | 0y<br>2.24 [2.22, 2.26]<br>5y<br>2.17 [2.15, 2.20] | 0y<br>2.25 [2.22, 2.27]<br>5y<br>2.16 [2.13, 2.18] | 0y<br>2.25 [2.23, 2.28]<br>5y<br>2.14 [2.11, 2.16] | 0y<br>2.28 [2.24, 2.32]<br>5y<br>2.09 [2.04, 2.13] |
| **75th<br>0.9** | 0y<br>2.29 [2.25, 2.32]<br>5y<br>2.29 [2.25, 2.32] | 0y<br>2.31 [2.28, 2.33]<br>5y<br>2.28 [2.25, 2.30] | 0y<br>2.32 [2.30, 2.35]<br>5y<br>2.27 [2.25, 2.30] | 0y<br>2.34 [2.32, 2.37]<br>5y<br>2.27 [2.24, 2.30] | 0y<br>2.39 [2.34, 2.43]<br>5y<br>2.25 [2.20, 2.30] |
| **99th<br>7.3** | 0y<br>2.46 [2.40, 2.52]<br>5y<br>2.56 [2.49, 2.63] | 0y<br>2.52 [2.48, 2.56]<br>5y<br>2.61 [2.56, 2.65] | 0y<br>2.56 [2.53, 2.59]<br>5y<br>2.63 [2.60, 2.67] | 0y<br>2.61 [2.57, 2.65]<br>5y<br>2.67 [2.62, 2.71] | 0y<br>2.73 [2.64, 2.81]<br>5y<br>2.76 [2.66, 2.86] |

**Supplementary Table 5A. Model-Predicted Cartilage Thickness With 95% Confidence Intervals: Medial Femur.** Primary-model predictions of mean cartilage thickness (mm) are shown at baseline and five years, at mean age, male sex, and baseline KL grade 0. Rows and columns represent the 1st, 25th, 50th, 75th, and 99th percentiles of limb-level HKA and participant-level BMI, respectively; displayed rounded values are used for prediction. Each cell shows baseline (0y) on the first line and five years (5y) on the second, with pointwise 95% confidence intervals in brackets beside each predicted mean, calculated from the fixed-effect covariance matrix. These describe uncertainty in the reference-profile mean, not variation between individual limbs. These estimates provide uncertainty around the medial trajectories illustrated in main manuscript Figure 3.

| HKA Percentile<br>Angle (°) | BMI 1st<br>20 kg/m² | BMI 25th<br>25 kg/m² | BMI 50th<br>28 kg/m² | BMI 75th<br>32 kg/m² | BMI 99th<br>42 kg/m² |
|---|---|---|---|---|---|
| **1st**<br>**-9.6** | 0y<br>1.91 [1.87, 1.96]<br>5y<br>1.79 [1.74, 1.84] | 0y<br>1.91 [1.88, 1.94]<br>5y<br>1.76 [1.73, 1.79] | 0y<br>1.91 [1.88, 1.93]<br>5y<br>1.74 [1.71, 1.77] | 0y<br>1.91 [1.88, 1.94]<br>5y<br>1.71 [1.68, 1.74] | 0y<br>1.90 [1.84, 1.96]<br>5y<br>1.65 [1.58, 1.72] |
| **25th**<br>**-3.4** | 0y<br>2.00 [1.97, 2.02]<br>5y<br>1.92 [1.89, 1.95] | 0y<br>2.01 [1.99, 2.03]<br>5y<br>1.93 [1.90, 1.95] | 0y<br>2.02 [2.00, 2.04]<br>5y<br>1.93 [1.91, 1.95] | 0y<br>2.03 [2.01, 2.05]<br>5y<br>1.93 [1.91, 1.95] | 0y<br>2.06 [2.02, 2.10]<br>5y<br>1.93 [1.89, 1.97] |
| **50th**<br>**-1.2** | 0y<br>2.03 [2.00, 2.05]<br>5y<br>1.97 [1.94, 2.00] | 0y<br>2.05 [2.03, 2.07]<br>5y<br>1.99 [1.96, 2.01] | 0y<br>2.06 [2.04, 2.08]<br>5y<br>1.99 [1.98, 2.01] | 0y<br>2.08 [2.06, 2.10]<br>5y<br>2.01 [1.99, 2.03] | 0y<br>2.12 [2.08, 2.15]<br>5y<br>2.04 [2.00, 2.07] |
| **75th**<br>**0.9** | 0y<br>2.05 [2.03, 2.08]<br>5y<br>2.02 [1.99, 2.04] | 0y<br>2.08 [2.06, 2.10]<br>5y<br>2.04 [2.02, 2.06] | 0y<br>2.10 [2.08, 2.12]<br>5y<br>2.06 [2.04, 2.08] | 0y<br>2.12 [2.10, 2.14]<br>5y<br>2.08 [2.06, 2.10] | 0y<br>2.17 [2.13, 2.21]<br>5y<br>2.13 [2.09, 2.17] |
| **99th**<br>**7.3** | 0y<br>2.14 [2.09, 2.19]<br>5y<br>2.15 [2.10, 2.20] | 0y<br>2.19 [2.15, 2.22]<br>5y<br>2.22 [2.18, 2.25] | 0y<br>2.21 [2.19, 2.24]<br>5y<br>2.25 [2.22, 2.28] | 0y<br>2.25 [2.22, 2.28]<br>5y<br>2.30 [2.27, 2.34] | 0y<br>2.34 [2.27, 2.40]<br>5y<br>2.43 [2.36, 2.50] |

**Supplementary Table 5B. Model-Predicted Cartilage Thickness With 95% Confidence Intervals: Medial Tibia.** Primary-model predictions of mean cartilage thickness (mm) are shown at baseline and five years, at mean age, male sex, and baseline KL grade 0. Rows and columns represent the 1st, 25th, 50th, 75th, and 99th percentiles of limb-level HKA and participant-level BMI, respectively; displayed rounded values are used for prediction. Each cell shows baseline (0y) on the first line and five years (5y) on the second, with pointwise 95% confidence intervals in brackets beside each predicted mean, calculated from the fixed-effect covariance matrix. These describe uncertainty in the reference-profile mean, not variation between individual limbs. These estimates provide uncertainty around the medial trajectories illustrated in main manuscript Figure 3.

| HKA Percentile<br>Angle (°) | BMI 1st<br>20 kg/m² | BMI 25th<br>25 kg/m² | BMI 50th<br>28 kg/m² | BMI 75th<br>32 kg/m² | BMI 99th<br>42 kg/m² |
|---|---|---|---|---|---|
| **1st<br>-9.6** | 0y<br>2.15 [2.09, 2.20]<br>5y<br>2.09 [2.04, 2.15] | 0y<br>2.15 [2.11, 2.19]<br>5y<br>2.09 [2.05, 2.13] | 0y<br>2.15 [2.12, 2.19]<br>5y<br>2.09 [2.05, 2.13] | 0y<br>2.15 [2.12, 2.19]<br>5y<br>2.09 [2.05, 2.13] | 0y<br>2.16 [2.09, 2.22]<br>5y<br>2.09 [2.02, 2.16] |
| **25th<br>-3.4** | 0y<br>2.10 [2.07, 2.13]<br>5y<br>2.11 [2.08, 2.14] | 0y<br>2.12 [2.10, 2.14]<br>5y<br>2.13 [2.11, 2.15] | 0y<br>2.14 [2.12, 2.16]<br>5y<br>2.14 [2.12, 2.16] | 0y<br>2.15 [2.13, 2.18]<br>5y<br>2.16 [2.14, 2.18] | 0y<br>2.20 [2.16, 2.24]<br>5y<br>2.20 [2.16, 2.24] |
| **50th<br>-1.2** | 0y<br>2.08 [2.05, 2.10]<br>5y<br>2.09 [2.06, 2.11] | 0y<br>2.11 [2.09, 2.13]<br>5y<br>2.11 [2.09, 2.13] | 0y<br>2.13 [2.11, 2.14]<br>5y<br>2.13 [2.11, 2.15] | 0y<br>2.15 [2.13, 2.17]<br>5y<br>2.15 [2.13, 2.17] | 0y<br>2.21 [2.17, 2.25]<br>5y<br>2.21 [2.17, 2.25] |
| **75th<br>0.9** | 0y<br>2.05 [2.02, 2.08]<br>5y<br>2.04 [2.02, 2.07] | 0y<br>2.09 [2.07, 2.11]<br>5y<br>2.08 [2.06, 2.10] | 0y<br>2.11 [2.09, 2.13]<br>5y<br>2.10 [2.08, 2.12] | 0y<br>2.14 [2.12, 2.16]<br>5y<br>2.13 [2.11, 2.15] | 0y<br>2.22 [2.18, 2.26]<br>5y<br>2.20 [2.16, 2.24] |
| **99th<br>7.3** | 0y<br>1.96 [1.90, 2.01]<br>5y<br>1.82 [1.77, 1.88] | 0y<br>2.01 [1.98, 2.05]<br>5y<br>1.88 [1.84, 1.92] | 0y<br>2.05 [2.02, 2.08]<br>5y<br>1.92 [1.88, 1.95] | 0y<br>2.10 [2.06, 2.14]<br>5y<br>1.96 [1.92, 2.00] | 0y<br>2.22 [2.14, 2.29]<br>5y<br>2.07 [2.00, 2.15] |

**Supplementary Table 5C. Model-Predicted Cartilage Thickness With 95% Confidence Intervals: Lateral Femur.** Primary-model predictions of mean cartilage thickness (mm) are shown at baseline and five years, at mean age, male sex, and baseline KL grade 0. Rows and columns represent the 1st, 25th, 50th, 75th, and 99th percentiles of limb-level HKA and participant-level BMI, respectively; displayed rounded values are used for prediction. Each cell shows baseline (0y) on the first line and five years (5y) on the second, with pointwise 95% confidence intervals in brackets beside each predicted mean, calculated from the fixed-effect covariance matrix. These describe uncertainty in the reference-profile mean, not variation between individual limbs. These estimates extend the reference-profile predictions to the lateral region.

| HKA Percentile<br>Angle (°) | BMI 1st<br>20 kg/m² | BMI 25th<br>25 kg/m² | BMI 50th<br>28 kg/m² | BMI 75th<br>32 kg/m² | BMI 99th<br>42 kg/m² |
|---|---|---|---|---|---|
| **1st<br>-9.6** | 0y<br>2.95 [2.87, 3.03]<br>5y<br>2.83 [2.75, 2.91] | 0y<br>2.96 [2.90, 3.02]<br>5y<br>2.84 [2.78, 2.90] | 0y<br>2.97 [2.92, 3.02]<br>5y<br>2.85 [2.79, 2.90] | 0y<br>2.97 [2.92, 3.03]<br>5y<br>2.86 [2.80, 2.92] | 0y<br>2.99 [2.90, 3.09]<br>5y<br>2.88 [2.78, 2.98] |
| **25th<br>-3.4** | 0y<br>2.75 [2.71, 2.79]<br>5y<br>2.66 [2.62, 2.70] | 0y<br>2.77 [2.74, 2.80]<br>5y<br>2.68 [2.65, 2.71] | 0y<br>2.78 [2.75, 2.81]<br>5y<br>2.70 [2.67, 2.72] | 0y<br>2.80 [2.77, 2.83]<br>5y<br>2.71 [2.68, 2.74] | 0y<br>2.84 [2.78, 2.89]<br>5y<br>2.75 [2.70, 2.81] |
| **50th<br>-1.2** | 0y<br>2.67 [2.63, 2.70]<br>5y<br>2.56 [2.52, 2.60] | 0y<br>2.69 [2.66, 2.72]<br>5y<br>2.59 [2.56, 2.62] | 0y<br>2.70 [2.68, 2.73]<br>5y<br>2.60 [2.58, 2.63] | 0y<br>2.72 [2.69, 2.75]<br>5y<br>2.62 [2.59, 2.65] | 0y<br>2.77 [2.71, 2.82]<br>5y<br>2.67 [2.61, 2.73] |
| **75th<br>0.9** | 0y<br>2.58 [2.54, 2.62]<br>5y<br>2.45 [2.41, 2.49] | 0y<br>2.61 [2.58, 2.64]<br>5y<br>2.48 [2.45, 2.51] | 0y<br>2.62 [2.59, 2.65]<br>5y<br>2.49 [2.47, 2.52] | 0y<br>2.64 [2.61, 2.67]<br>5y<br>2.52 [2.48, 2.55] | 0y<br>2.70 [2.64, 2.75]<br>5y<br>2.57 [2.51, 2.63] |
| **99th<br>7.3** | 0y<br>2.28 [2.21, 2.36]<br>5y<br>1.99 [1.91, 2.07] | 0y<br>2.32 [2.27, 2.37]<br>5y<br>2.03 [1.97, 2.09] | 0y<br>2.34 [2.29, 2.39]<br>5y<br>2.05 [1.99, 2.11] | 0y<br>2.37 [2.31, 2.43]<br>5y<br>2.08 [2.02, 2.14] | 0y<br>2.44 [2.34, 2.55]<br>5y<br>2.16 [2.05, 2.26] |

**Supplementary Table 5D. Model-Predicted Cartilage Thickness With 95% Confidence Intervals: Lateral Tibia.** Primary-model predictions of mean cartilage thickness (mm) are shown at baseline and five years, at mean age, male sex, and baseline KL grade 0. Rows and columns represent the 1st, 25th, 50th, 75th, and 99th percentiles of limb-level HKA and participant-level BMI, respectively; displayed rounded values are used for prediction. Each cell shows baseline (0y) on the first line and five years (5y) on the second, with pointwise 95% confidence intervals in brackets beside each predicted mean, calculated from the fixed-effect covariance matrix. These describe uncertainty in the reference-profile mean, not variation between individual limbs. These estimates extend the reference-profile predictions to the lateral region.

| Region | Effect | Raw P | BH-FDR P | FDR < 0.05 |
|---|---|---|---|---|
| Medial femur | time x BMI x HKA | 0.042 | 0.042 | Yes |
| Medial tibia | time x BMI x HKA | <0.001 | <0.001 | Yes |
| Lateral femur | time x $HKA^2$ | <0.001 | <0.001 | Yes |
| Lateral tibia | time x $HKA^2$ | <0.001 | <0.001 | Yes |

**Supplementary Table 6. Benjamini-Hochberg False Discovery Rate Correction.** The correction was applied once across the four primary interaction tests: time x BMI x HKA in the medial femur and medial tibia, and time x $HKA^2$ in the lateral femur and lateral tibia. All four remained significant after correction.

**A. Time linearity**

| Region | Primary interaction | Linear estimate (95% CI) | Flexible estimate (95% CI) | Change in coefficient magnitude (%) |
|---|---|---|---|---|
| Medial Femur | Time x HKA x BMI | +0.0016 (+0.0001, +0.0031) | +0.0016 (+0.0001, +0.0031) | 0.78 |
| Medial Tibia | Time x HKA x BMI | +0.0018 (+0.0008, +0.0028) | +0.0018 (+0.0009, +0.0028) | 0.52 |
| Lateral Femur | Time x $HKA^2$ | -0.0032 (-0.0038, -0.0027) | -0.0032 (-0.0038, -0.0027) | -0.40 |
| Lateral Tibia | Time x $HKA^2$ | -0.0033 (-0.0040, -0.0026) | -0.0033 (-0.0039, -0.0026) | -0.81 |

**B. HKA linearity**

| Region | Linear mean absolute difference (mm/year) | Quadratic mean absolute difference (mm/year) | Natural spline mean absolute difference (mm/year) | Selected form |
|---|---|---|---|---|
| Medial Femur | 0.0045 | 0.0041 | 0.0040 | linear |
| Medial Tibia | 0.0028 | 0.0016 | 0.0013 | linear |
| Lateral Femur | 0.0038 | 0.0014 | 0.0019 | quadratic |
| Lateral Tibia | 0.0057 | 0.0023 | 0.0016 | quadratic |

**C. BMI linearity**

| Region | Linear mean absolute difference (mm/year) | Quadratic mean absolute difference (mm/year) | Absolute improvement (mm/year) | Time x $BMI^2$ coefficient | Time x $BMI^2$ p value |
|---|---|---|---|---|---|
| Medial Femur | 0.0058 | 0.0044 | 0.0014 | 0.0021 | <0.001 |
| Medial Tibia | 0.0037 | 0.0029 | 0.0008 | 0.0014 | <0.001 |
| Lateral Femur | 0.0042 | 0.0048 | -0.0006 | 0.0006 | 0.093 |
| Lateral Tibia | 0.0040 | 0.0038 | 0.0001 | 0.0007 | 0.094 |

**Supplementary Table 7. Evaluation of the Linearity Assumptions for Time, HKA, and BMI.** (A) Allowing a flexible common time trajectory changed primary interaction estimates by less than 1%, supporting linear time for estimating average annual differences in thickness change. (B) Linear, quadratic, and natural-spline HKA models were compared with limb-level rates. Nonlinear forms improved agreement, particularly in the medial tibia and lateral regions. The medial tibial interaction sensitivity is reported separately in Supplementary Table 8. (C) Quadratic BMI improved agreement medially, with little or inconsistent improvement laterally. Primary models retained linear medial HKA and BMI terms to summarize average exposure-associated differences in thinning rate, and quadratic lateral HKA terms to represent faster thinning toward both alignment extremes. Mean absolute differences are reported in mm/year, with smaller values indicating closer agreement with limb-level rates.

| Term | Linear β [95% CI] | p value | Quadratic β [95% CI] | p value |
|---|---|---|---|---|
| BMI x HKA x time | 0.0018 [0.0008, 0.0028] | <0.001 | 0.0015 [0.0005, 0.0025] | 0.003 |
| $HKA^2$ x time | Not included | — | −0.0019 [−0.0025, −0.0013] | <0.001 |

**Supplementary Table 8. Medial Tibial BMI–Alignment Interaction with Linear and Quadratic HKA.** Models used identical observations, scanner correction, covariates, random effects, and BMI and HKA standardization. The quadratic model added only $HKA^2$ and time × $HKA^2$, retaining all existing terms. The time × BMI × HKA interaction remained positive and significant in both models before and after four-test FDR adjustment (linear: $p<0.001$; quadratic: $p=0.003$). At mean BMI, the mean absolute difference between models in predicted five-year cartilage thickness change was 0.0095 mm across all baseline limbs, approximately 41-42% smaller than the corresponding differences in the lateral femur (0.0162 mm) and lateral tibia (0.0165 mm). Given that medial tibial predictions were less sensitive to the inclusion of $HKA^2$ and time × $HKA^2$ than lateral predictions, and the primary interaction (time × HKA × BMI) remained significant, we retained the linear HKA specification.

| Region | Effect (interaction term) | A. Primary: KL, all visits β / 95% CI / p | B. KL, follow-up sample β / 95% CI / p | C. Baseline thickness replaces KL β / 95% CI / p |
|---|---|---|---|---|
| Medial femur | BMI x HKA x time | +0.0016 [+0.0001, +0.0031] $p=0.042$ | +0.0019 [+0.0001, +0.0038] $p=0.043$ | +0.0019 [+0.0000, +0.0037] $p=0.047$ |
| | HKA x time | +0.0132 [+0.0117, +0.0146] $p<0.001$ | +0.0126 [+0.0108, +0.0143] $p<0.001$ | +0.0129 [+0.0112, +0.0147] $p<0.001$ |
| | BMI x time | -0.0070 [-0.0084, -0.0055] $p<0.001$ | -0.0076 [-0.0093, -0.0058] $p<0.001$ | -0.0077 [-0.0095, -0.0059] $p<0.001$ |
| Medial tibia | BMI x HKA x time | +0.0018 [+0.0008, +0.0028] $p<0.001$ | +0.0026 [+0.0013, +0.0039] $p<0.001$ | +0.0028 [+0.0015, +0.0040] $p<0.001$ |
| | HKA x time | +0.0087 [+0.0078, +0.0096] $p<0.001$ | +0.0082 [+0.0070, +0.0094] $p<0.001$ | +0.0082 [+0.0070, +0.0094] $p<0.001$ |
| | BMI x time | -0.0012 [-0.0021, -0.0002] $p=0.014$ | -0.0019 [-0.0031, -0.0007] $p=0.002$ | -0.0018 [-0.0030, -0.0007] $p=0.002$ |
| Lateral femur | HKA x time | -0.0028 [-0.0037, -0.0019] $p<0.001$ | -0.0027 [-0.0038, -0.0016] $p<0.001$ | -0.0026 [-0.0037, -0.0015] $p<0.001$ |
| | $HKA^2$ x time | -0.0032 [-0.0038, -0.0027] $p<0.001$ | -0.0033 [-0.0040, -0.0026] $p<0.001$ | -0.0031 [-0.0038, -0.0024] $p<0.001$ |
| | BMI x time | -0.0005 [-0.0014, +0.0004] $p=0.242$ | -0.0011 [-0.0023, +0.0000] $p=0.056$ | -0.0010 [-0.0021, +0.0001] $p=0.087$ |
| Lateral tibia | HKA x time | -0.0067 [-0.0078, -0.0056] $p<0.001$ | -0.0068 [-0.0082, -0.0054] $p<0.001$ | -0.0070 [-0.0084, -0.0055] $p<0.001$ |
| | $HKA^2$ x time | -0.0033 [-0.0040, -0.0026] $p<0.001$ | -0.0037 [-0.0046, -0.0028] $p<0.001$ | -0.0037 [-0.0045, -0.0028] $p<0.001$ |
| | BMI x time | +0.0002 [-0.0010, +0.0013] $p=0.757$ | -0.0003 [-0.0017, +0.0012] $p=0.706$ | -0.0001 [-0.0015, +0.0014] $p=0.917$ |

**Supplementary Table 9A. Sensitivity Analysis Replacing KL Grade with Baseline Cartilage Thickness.** Column A shows the primary KL-adjusted model using all visits. Because baseline cartilage thickness is also the outcome at time zero, Column B repeats the KL-adjusted model using follow-up observations only. Column C replaces KL grade with baseline compartment-specific cartilage thickness using the same follow-up observations as Column B. Comparing Columns A and B shows the effect of removing the baseline outcome, whereas comparing Columns B and C isolates the effect of replacing KL grade with baseline cartilage thickness. Across the four regions, the primary estimates retained the same direction and statistical significance and changed by no more than 6% between Columns B and C, including the medial time x BMI x HKA interactions (femur: 0.0019 vs 0.0019; tibia: 0.0026 vs 0.0028) and lateral time x $HKA^2$ interactions (femur: −0.0033 vs −0.0031; tibia: −0.0037 vs −0.0037). The findings were consistent whether baseline disease severity was represented by KL grade or baseline cartilage thickness.

| Region | Effect (interaction term) | A. Primary: KL grade β / 95% CI / p | B. Baseline JSN grade replaces KL β / 95% CI / p |
|---|---|---|---|
| Medial femur | BMI x HKA x time | +0.0016 [+0.0001, +0.0031] p=0.042 | +0.0016 [+0.0001, +0.0031] p=0.039 |
| | HKA x time | +0.0132 [+0.0117, +0.0146] p<0.001 | +0.0131 [+0.0117, +0.0145] p<0.001 |
| | BMI x time | -0.0070 [-0.0084, -0.0055] p<0.001 | -0.0070 [-0.0084, -0.0055] p<0.001 |
| Medial tibia | BMI x HKA x time | +0.0018 [+0.0008, +0.0028] p<0.001 | +0.0018 [+0.0008, +0.0028] p<0.001 |
| | HKA x time | +0.0087 [+0.0078, +0.0096] p<0.001 | +0.0087 [+0.0078, +0.0096] p<0.001 |
| | BMI x time | -0.0012 [-0.0021, -0.0002] p=0.014 | -0.0012 [-0.0021, -0.0002] p=0.013 |
| Lateral femur | HKA x time | -0.0028 [-0.0037, -0.0019] p<0.001 | -0.0028 [-0.0037, -0.0019] p<0.001 |
| | $HKA^2$ x time | -0.0032 [-0.0038, -0.0027] p<0.001 | -0.0032 [-0.0038, -0.0027] p<0.001 |
| | BMI x time | -0.0005 [-0.0014, +0.0004] p=0.242 | -0.0005 [-0.0014, +0.0004] p=0.242 |
| Lateral tibia | HKA x time | -0.0067 [-0.0078, -0.0056] p<0.001 | -0.0067 [-0.0078, -0.0055] p<0.001 |
| | $HKA^2$ x time | -0.0033 [-0.0040, -0.0026] p<0.001 | -0.0033 [-0.0040, -0.0026] p<0.001 |
| | BMI x time | +0.0002 [-0.0010, +0.0013] p=0.757 | +0.0002 [-0.0010, +0.0013] p=0.761 |

**Supplementary Table 9B. Sensitivity Analysis Replacing KL Grade with Baseline Compartment-specific Joint-Space-Narrowing Grade.** Column A shows the primary models adjusted for KL grade, and Column B replaces KL grade with baseline compartment-specific joint-space-narrowing grade using the same limbs and visits; all other model components were unchanged. Comparing Columns A and B therefore isolates the effect of representing baseline disease severity with compartment-specific narrowing rather than KL grade. The estimates were nearly identical, including the medial time x BMI x HKA interactions (femur: 0.0016 vs 0.0016; tibia: 0.0018 vs 0.0018) and lateral time x $HKA^2$ interactions (femur: −0.0032 vs −0.0032; tibia: −0.0033 vs −0.0033). Replacing KL grade with compartment-specific joint-space-narrowing grade did not change the direction, magnitude, or statistical significance of the findings.

| Region | Effect (interaction term) | A. Primary: KL, full cohort β / 95% CI / p | B. KL, JSW subset β / 95% CI / p | C. Quantitative JSW replaces KL β / 95% CI / p |
|---|---|---|---|---|
| Medial femur | BMI x HKA x time | +0.0016 [+0.0001, +0.0031] p=0.042 | +0.0006 [-0.0013, +0.0025] p=0.528 | +0.0006 [-0.0012, +0.0025] p=0.513 |
| | HKA x time | +0.0132 [+0.0117, +0.0146] p<0.001 | +0.0148 [+0.0130, +0.0166] p<0.001 | +0.0147 [+0.0130, +0.0165] p<0.001 |
| | BMI x time | -0.0070 [-0.0084, -0.0055] p<0.001 | -0.0062 [-0.0081, -0.0043] p<0.001 | -0.0063 [-0.0082, -0.0044] p<0.001 |
| Medial tibia | BMI x HKA x time | +0.0018 [+0.0008, +0.0028] p<0.001 | +0.0011 [-0.0001, +0.0022] p=0.072 | +0.0010 [-0.0001, +0.0022] p=0.084 |
| | HKA x time | +0.0087 [+0.0078, +0.0096] p<0.001 | +0.0100 [+0.0089, +0.0111] p<0.001 | +0.0099 [+0.0088, +0.0110] p<0.001 |
| | BMI x time | -0.0012 [-0.0021, -0.0002] p=0.014 | -0.0007 [-0.0019, +0.0005] p=0.244 | -0.0007 [-0.0019, +0.0004] p=0.224 |
| Lateral femur | HKA x time | -0.0028 [-0.0037, -0.0019] p<0.001 | -0.0041 [-0.0052, -0.0030] p<0.001 | -0.0040 [-0.0051, -0.0029] p<0.001 |
| | $HKA^2$ x time | -0.0032 [-0.0038, -0.0027] p<0.001 | -0.0035 [-0.0042, -0.0029] p<0.001 | -0.0035 [-0.0042, -0.0028] p<0.001 |
| | BMI x time | -0.0005 [-0.0014, +0.0004] p=0.242 | +0.0002 [-0.0010, +0.0013] p=0.793 | +0.0002 [-0.0010, +0.0013] p=0.789 |
| Lateral tibia | HKA x time | -0.0067 [-0.0078, -0.0056] p<0.001 | -0.0082 [-0.0096, -0.0068] p<0.001 | -0.0082 [-0.0096, -0.0068] p<0.001 |
| | $HKA^2$ x time | -0.0033 [-0.0040, -0.0026] p<0.001 | -0.0035 [-0.0044, -0.0027] p<0.001 | -0.0036 [-0.0044, -0.0027] p<0.001 |
| | BMI x time | +0.0002 [-0.0010, +0.0013] p=0.757 | +0.0011 [-0.0004, +0.0026] p=0.138 | +0.0011 [-0.0003, +0.0026] p=0.135 |

**Supplementary Table 9C. Sensitivity Analysis Replacing KL Grade with Baseline Quantitative Joint-Space Width.** Quantitative joint-space width was available for 4,209 limbs in the medial analysis and 4,159 limbs in the lateral analysis. Column A shows the primary KL-adjusted model in the full cohort. Column B repeats that model in the joint-space-width subset, and Column C replaces KL grade with baseline compartment-specific quantitative joint-space width in the same subset. Comparing Columns A and B shows the effect of restricting the sample, whereas comparing Columns B and C isolates the effect of replacing KL grade with joint-space width. Estimates in Columns B and C were nearly identical, including the medial time x BMI x HKA interactions (femur: 0.0006 vs 0.0006; tibia: 0.0011 vs 0.0010) and lateral time x $HKA^2$ interactions (femur: −0.0035 vs −0.0035; tibia: −0.0035 vs −0.0036). The medial interactions were attenuated and not significant in both subsets. Replacing KL grade with joint-space width did not change the direction, magnitude, or statistical significance of the findings within the same subset; the attenuation relative to the full cohort reflected subset selection rather than the joint-space-width measure.

| Region | Effect (interaction term) | A. Primary: full cohort β / 95% CI / p | B. Restricted to baseline JSN=0 β / 95% CI / p |
|---|---|---|---|
| Medial femur | BMI x HKA x time | +0.0016 [+0.0001, +0.0031] p=0.042 | +0.0013 [-0.0003, +0.0029] p=0.107 |
| | HKA x time | +0.0132 [+0.0117, +0.0146] p<0.001 | +0.0051 [+0.0036, +0.0066] p<0.001 |
| | BMI x time | -0.0070 [-0.0084, -0.0055] p<0.001 | -0.0046 [-0.0061, -0.0032] p<0.001 |
| Medial tibia | BMI x HKA x time | +0.0018 [+0.0008, +0.0028] p<0.001 | +0.0013 [+0.0002, +0.0023] p=0.019 |
| | HKA x time | +0.0087 [+0.0078, +0.0096] p<0.001 | +0.0019 [+0.0009, +0.0029] p<0.001 |
| | BMI x time | -0.0012 [-0.0021, -0.0002] p=0.014 | +0.0004 [-0.0005, +0.0014] p=0.375 |
| Lateral femur | HKA x time | -0.0028 [-0.0037, -0.0019] p<0.001 | +0.0018 [+0.0009, +0.0027] p<0.001 |
| | $HKA^2$ x time | -0.0032 [-0.0038, -0.0027] p<0.001 | -0.0005 [-0.0011, +0.0001] p=0.110 |
| | BMI x time | -0.0005 [-0.0014, +0.0004] p=0.242 | -0.0008 [-0.0016, -0.0000] p=0.041 |
| Lateral tibia | HKA x time | -0.0067 [-0.0078, -0.0056] p<0.001 | -0.0025 [-0.0036, -0.0013] p<0.001 |
| | $HKA^2$ x time | -0.0033 [-0.0040, -0.0026] p<0.001 | -0.0018 [-0.0026, -0.0011] p<0.001 |
| | BMI x time | +0.0002 [-0.0010, +0.0013] p=0.757 | +0.0000 [-0.0010, +0.0010] p=0.959 |

**Supplementary Table 9D. Sensitivity Analysis Restricted to Limbs Without Baseline Compartment-specific Narrowing.** Column A shows the primary model in the full cohort, and Column B shows the same model restricted to limbs with a baseline joint-space-narrowing grade of 0 in the analyzed region (medial, 3,658 limbs; lateral, 5,380 limbs). In the medial analysis, the restriction retained only 518 of 1,355 limbs with mild-to-severe varus alignment (38.2%). The medial tibia time x BMI x HKA interaction remained significant ($\beta = 0.0013$, $p = 0.019$), whereas the medial femur estimate remained positive but was no longer significant ($\beta = 0.0013$, $p = 0.107$). In the lateral analysis, the restriction removed 320 of 1,078 limbs with mild-to-severe valgus alignment (29.7%) but only 24 of 1,355 limbs with mild-to-severe varus alignment (1.8%), reducing representation of the valgus arm. The lateral tibial time x $HKA^2$ interaction remained significant ($\beta = -0.0018$, $p < 0.001$), whereas the lateral femoral estimate was attenuated and not significant ($\beta = -0.0005$, $p = 0.110$). The restriction preserved the direction of the primary findings but reduced samples at the relevant alignment extremes, which likely contributed to the attenuated results.

| Region | Cohort | Limbs | Participants | MRI | Estimate (95% CI) | p |
|---|---|---|---|---|---|---|
| Medial Femur | Full Cohort | 5,930 | 3,191 | 20,841 | 0.0016 [0.0001, 0.0031] | 0.042 |
| | Baseline KL = 0 | 2,010 | 1,381 | 6,796 | 0.0039 [0.0017, 0.0062] | <0.001 |
| Medial Tibia | Full Cohort | 5,930 | 3,191 | 20,843 | 0.0018 [0.0008, 0.0028] | <0.001 |
| | Baseline KL = 0 | 2,010 | 1,381 | 6,797 | 0.0024 [0.0008, 0.0040] | 0.003 |
| Lateral Femur | Full Cohort | 5,930 | 3,191 | 20,839 | -0.0032 [-0.0038, -0.0027] | <0.001 |
| | Baseline KL = 0 | 2,010 | 1,381 | 6,795 | 0.0010 [-0.0001, 0.0021] | 0.077 |
| Lateral Tibia | Full Cohort | 5,930 | 3,191 | 20,843 | -0.0033 [-0.0040, -0.0026] | <0.001 |
| | Baseline KL = 0 | 2,010 | 1,381 | 6,797 | -0.0025 [-0.0042, -0.0008] | 0.004 |

**Supplementary Table 9E. Sensitivity Analysis Restricted to Limbs with Baseline KL Grade 0.** Primary models were repeated in 2,010 limbs from 1,381 participants with KL grade 0 at baseline, retaining all eligible follow-up observations regardless of subsequent KL grade. This subgroup differs from the reference group required to have KL grade 0 at every included visit. Age and sex adjustment, random effects, and full-cohort centering and standardization were retained; KL was omitted from the restricted models because it was constant. Estimates represent the time x BMI x HKA interaction in the medial regions and the time x $HKA^2$ interaction in the lateral regions. Time is measured in years; BMI and HKA use full-cohort baseline SD units, and positive HKA denotes valgus. Confidence intervals are Wald 95% intervals; p values use Satterthwaite degrees of freedom and are unadjusted for this exploratory analysis. Both medial interactions remained significant (femur: $\beta = 0.0039$, $p < 0.001$; tibia: $\beta = 0.0024$, $p = 0.003$), supporting a compounding association in limbs without baseline radiographic OA. The lateral tibial estimate ($HKA^2$ x time) remained significant ($\beta = -0.0025$, $p = 0.004$); the lateral femoral estimate changed sign and was not significant ($\beta = 0.0010$, $p = 0.077$).

| Region | Effect (Interaction Term) | A. Baseline BMI β / 95% CI / p | B. Visit-Specific BMI β / 95% CI / p | C. Mean Follow-Up BMI β / 95% CI / p |
|---|---|---|---|---|
| Medial femur | time x BMI x HKA | +0.0016 [+0.0001, +0.0031] p=0.042 | +0.0016 [+0.0002, +0.0030] p=0.025 | +0.0022 [+0.0007, +0.0037] p=0.004 |
| | time x HKA | +0.0132 [+0.0117, +0.0146] p<0.001 | +0.0130 [+0.0116, +0.0145] p<0.001 | +0.0131 [+0.0117, +0.0145] p<0.001 |
| | time x BMI | -0.0070 [-0.0084, -0.0055] p<0.001 | -0.0075 [-0.0088, -0.0061] p<0.001 | -0.0077 [-0.0092, -0.0063] p<0.001 |
| Medial tibia | time x BMI x HKA | +0.0018 [+0.0008, +0.0028] p<0.001 | +0.0017 [+0.0008, +0.0026] p<0.001 | +0.0020 [+0.0010, +0.0030] p<0.001 |
| | time x HKA | +0.0087 [+0.0078, +0.0096] p<0.001 | +0.0087 [+0.0078, +0.0096] p<0.001 | +0.0087 [+0.0077, +0.0096] p<0.001 |
| | time x BMI | -0.0012 [-0.0021, -0.0002] p=0.014 | -0.0016 [-0.0025, -0.0007] p<0.001 | -0.0014 [-0.0024, -0.0005] p=0.002 |
| Lateral femur | time x HKA | -0.0028 [-0.0037, -0.0019] p<0.001 | -0.0027 [-0.0036, -0.0018] p<0.001 | -0.0028 [-0.0037, -0.0019] p<0.001 |
| | time x $HKA^2$ | -0.0032 [-0.0038, -0.0027] p<0.001 | -0.0032 [-0.0038, -0.0027] p<0.001 | -0.0032 [-0.0038, -0.0027] p<0.001 |
| | time x BMI | -0.0005 [-0.0014, +0.0004] p=0.242 | -0.0009 [-0.0018, -0.0000] p=0.040 | -0.0006 [-0.0015, +0.0003] p=0.158 |
| Lateral tibia | time x HKA | -0.0067 [-0.0078, -0.0056] p<0.001 | -0.0067 [-0.0078, -0.0056] p<0.001 | -0.0067 [-0.0079, -0.0056] p<0.001 |
| | time x $HKA^2$ | -0.0033 [-0.0040, -0.0026] p<0.001 | -0.0033 [-0.0040, -0.0026] p<0.001 | -0.0033 [-0.0040, -0.0026] p<0.001 |
| | time x BMI | +0.0002 [-0.0010, +0.0013] p=0.757 | -0.0004 [-0.0015, +0.0007] p=0.446 | -0.0001 [-0.0012, +0.0011] p=0.877 |

**Supplementary Table 10. Sensitivity Analysis Using Follow-Up and Average BMI Measurements.** Column A represents the primary model using baseline BMI. Column B replaces baseline BMI with BMI measured at each visit, and Column C uses average BMI across each limb's observed follow-up. Comparing Columns B and C with Column A assesses whether incorporating BMI after baseline changes the findings. The medial time x BMI x HKA interactions remained positive and statistically significant across Columns A, B, and C. The lateral time x $HKA^2$ estimates remained negative and statistically significant across all three BMI definitions. Incorporating observed BMI during follow-up did not alter the primary malalignment or compounding findings.

| Region | Effect (Interaction Term) | A. Primary β / 95% CI / p | B. Baseline PASE Added β / 95% CI / p |
|---|---|---|---|
| Medial femur | time x BMI x HKA | +0.0016 [+0.0001, +0.0031] p=0.042 | +0.0016 [+0.0001, +0.0031] p=0.040 |
| | time x HKA | +0.0132 [+0.0117, +0.0146] p<0.001 | +0.0133 [+0.0119, +0.0148] p<0.001 |
| | time x BMI | -0.0070 [-0.0084, -0.0055] p<0.001 | -0.0068 [-0.0083, -0.0054] p<0.001 |
| | time x PASE | — | +0.0006 [-0.0008, +0.0021] p=0.374 |
| Medial tibia | time x BMI x HKA | +0.0018 [+0.0008, +0.0028] p<0.001 | +0.0018 [+0.0008, +0.0028] p<0.001 |
| | time x HKA | +0.0087 [+0.0078, +0.0096] p<0.001 | +0.0088 [+0.0079, +0.0098] p<0.001 |
| | time x BMI | -0.0012 [-0.0021, -0.0002] p=0.014 | -0.0011 [-0.0020, -0.0001] p=0.027 |
| | time x PASE | — | +0.0009 [-0.0000, +0.0018] p=0.061 |
| Lateral femur | time x HKA | -0.0028 [-0.0037, -0.0019] p<0.001 | -0.0027 [-0.0036, -0.0018] p<0.001 |
| | time x $HKA^2$ | -0.0032 [-0.0038, -0.0027] p<0.001 | -0.0032 [-0.0037, -0.0026] p<0.001 |
| | time x BMI | -0.0005 [-0.0014, +0.0004] p=0.242 | -0.0005 [-0.0014, +0.0004] p=0.242 |
| | time x PASE | — | -0.0001 [-0.0010, +0.0008] p=0.840 |
| Lateral tibia | time x HKA | -0.0067 [-0.0078, -0.0056] p<0.001 | -0.0067 [-0.0078, -0.0055] p<0.001 |
| | time x $HKA^2$ | -0.0033 [-0.0040, -0.0026] p<0.001 | -0.0032 [-0.0039, -0.0025] p<0.001 |
| | time x BMI | +0.0002 [-0.0010, +0.0013] p=0.757 | +0.0002 [-0.0009, +0.0014] p=0.721 |
| | time x PASE | — | -0.0004 [-0.0015, +0.0007] p=0.482 |

**Supplementary Table 11. Sensitivity Analysis Adjusting for Baseline Physical Activity.** Column A is the primary model. Column B adds baseline Physical Activity Scale for the Elderly (PASE) score and its interaction with time, allowing baseline self-reported activity to be associated with the subsequent rate of cartilage thickness change. Baseline PASE was available for 5,883 of 5,930 limbs. Comparing Columns A and B assesses the stability of the primary estimates after PASE adjustment in limbs with available measurements. The medial time x BMI x HKA estimate changes were negligible in the medial femur and medial tibia; the lateral time x $HKA^2$ estimate changes were also negligible. These associations retained their direction and statistical significance, with modest change in magnitude. The time x PASE interaction was not statistically significant in any region ($p = 0.061–0.840$). Adjustment for baseline self-reported physical activity did not alter the primary findings.

| Medial Femur | | | |
|---|---|---|---|
| **Term** | **β** | **95% CI** | **p** |
| Intercept | +2.2045 | [+2.1825, +2.2265] | <0.001 |
| **Main effects** | | | |
| Time (years) | -0.0189 | [-0.0203, -0.0175] | <0.001 |
| Body Mass (per SD) | +0.0137 | [-0.0000, +0.0274] | 0.050 |
| HKA angle (per SD) | +0.1474 | [+0.1377, +0.1571] | <0.001 |
| Age (per SD) | -0.0305 | [-0.0426, -0.0183] | <0.001 |
| Sex (female vs male) | -0.4271 | [-0.4540, -0.4002] | <0.001 |
| **KL grade** | | | |
| KL 1 (vs KL 0) | +0.0213 | [-0.0029, +0.0456] | 0.085 |
| KL 2 (vs KL 0) | +0.0182 | [-0.0044, +0.0408] | 0.114 |
| KL 3 (vs KL 0) | -0.2684 | [-0.2953, -0.2416] | <0.001 |
| KL 4 (vs KL 0) | -0.7632 | [-0.8029, -0.7236] | <0.001 |
| **Interactions** | | | |
| Time x Body Mass | -0.0072 | [-0.0086, -0.0057] | <0.001 |
| Time x HKA angle | +0.0128 | [+0.0114, +0.0143] | <0.001 |
| Body Mass x HKA angle | +0.0296 | [+0.0201, +0.0391] | <0.001 |
| Time x Body Mass x HKA angle | +0.0021 | [+0.0007, +0.0036] | 0.004 |
| **Medial Tibia** | | | |
| **Term** | **β** | **95% CI** | **p** |
| Intercept | +2.0239 | [+2.0050, +2.0427] | <0.001 |
| **Main effects** | | | |
| Time (years) | -0.0134 | [-0.0144, -0.0125] | <0.001 |
| Body Mass (per SD) | +0.0319 | [+0.0197, +0.0440] | <0.001 |
| HKA angle (per SD) | +0.0748 | [+0.0676, +0.0820] | <0.001 |
| Age (per SD) | -0.0375 | [-0.0484, -0.0267] | <0.001 |
| Sex (female vs male) | -0.2677 | [-0.2917, -0.2438] | <0.001 |
| **KL grade** | | | |
| KL 1 (vs KL 0) | +0.0238 | [+0.0065, +0.0412] | 0.007 |
| KL 2 (vs KL 0) | +0.0489 | [+0.0324, +0.0654] | <0.001 |
| KL 3 (vs KL 0) | -0.0495 | [-0.0691, -0.0300] | <0.001 |
| KL 4 (vs KL 0) | -0.3456 | [-0.3736, -0.3176] | <0.001 |
| **Interactions** | | | |
| Time x Body Mass | -0.0020 | [-0.0030, -0.0011] | <0.001 |
| Time x HKA angle | +0.0086 | [+0.0077, +0.0095] | <0.001 |
| Body Mass x HKA angle | +0.0180 | [+0.0110, +0.0251] | <0.001 |
| Time x Body Mass x HKA angle | +0.0014 | [+0.0005, +0.0024] | 0.004 |

**Supplementary Table 12A. Sensitivity Analysis Replacing BMI With Body Mass in Medial Regions.** Baseline body mass replaces BMI in the primary models, with all other model terms retained. Coefficients involving body mass or HKA are reported per 1-SD difference in the corresponding variable. The time x body mass x HKA interaction was positive and significant in the medial femur (β = 0.0021 mm/year per 1-SD body mass * 1-SD HKA, 95% CI [0.0007, 0.0036], p = 0.004) and medial tibia (β = 0.0014, 95% CI [0.0005, 0.0024], p = 0.004). The medial compounding interaction was therefore retained when body mass replaced BMI.

| Lateral Femur | | | |
|---|---|---|---|
| **Term** | **β** | **95% CI** | **p** |
| Intercept | +2.1041 | [+2.0851, +2.1231] | <0.001 |
| **Main effects** | | | |
| Time (years) | +0.0010 | [-0.0001, +0.0020] | 0.069 |
| Body Mass (per SD) | +0.0580 | [+0.0460, +0.0699] | <0.001 |
| HKA angle (per SD) | -0.0243 | [-0.0320, -0.0167] | <0.001 |
| $HKA^2$ (per $SD^2$) | -0.0093 | [-0.0132, -0.0053] | <0.001 |
| Age (per SD) | -0.0055 | [-0.0161, +0.0052] | 0.313 |
| Sex (female vs male) | -0.2459 | [-0.2695, -0.2223] | <0.001 |
| **KL grade** | | | |
| KL 1 (vs KL 0) | +0.0272 | [+0.0087, +0.0458] | 0.004 |
| KL 2 (vs KL 0) | +0.0745 | [+0.0569, +0.0920] | <0.001 |
| KL 3 (vs KL 0) | +0.0414 | [+0.0203, +0.0626] | <0.001 |
| KL 4 (vs KL 0) | -0.0362 | [-0.0690, -0.0033] | 0.031 |
| **Interactions** | | | |
| Time x Body Mass | -0.0032 | [-0.0041, -0.0023] | <0.001 |
| Time x HKA angle | -0.0030 | [-0.0039, -0.0021] | <0.001 |
| Time x $HKA^2$ | -0.0032 | [-0.0037, -0.0026] | <0.001 |
| Body Mass x HKA angle | +0.0055 | [-0.0019, +0.0129] | 0.145 |
| **Lateral Tibia** | | | |
| **Term** | **β** | **95% CI** | **p** |
| Intercept | +2.6518 | [+2.6241, +2.6795] | <0.001 |
| **Main effects** | | | |
| Time (years) | -0.0198 | [-0.0211, -0.0185] | <0.001 |
| Body Mass (per SD) | +0.0474 | [+0.0298, +0.0650] | <0.001 |
| HKA angle (per SD) | -0.1355 | [-0.1465, -0.1244] | <0.001 |
| $HKA^2$ (per $SD^2$) | -0.0133 | [-0.0190, -0.0077] | <0.001 |
| Age (per SD) | -0.0728 | [-0.0884, -0.0572] | <0.001 |
| Sex (female vs male) | -0.3391 | [-0.3736, -0.3045] | <0.001 |
| **KL grade** | | | |
| KL 1 (vs KL 0) | -0.0309 | [-0.0575, -0.0043] | 0.023 |
| KL 2 (vs KL 0) | -0.0750 | [-0.1002, -0.0498] | <0.001 |
| KL 3 (vs KL 0) | -0.2830 | [-0.3134, -0.2527] | <0.001 |
| KL 4 (vs KL 0) | -0.6038 | [-0.6508, -0.5567] | <0.001 |
| **Interactions** | | | |
| Time x Body Mass | -0.0021 | [-0.0032, -0.0009] | <0.001 |
| Time x HKA angle | -0.0069 | [-0.0080, -0.0057] | <0.001 |
| Time x $HKA^2$ | -0.0032 | [-0.0039, -0.0026] | <0.001 |
| Body Mass x HKA angle | -0.0026 | [-0.0133, +0.0080] | 0.629 |

**Supplementary Table 12B. Sensitivity Analysis Replacing BMI With Body Mass in Lateral Regions.** Baseline body mass replaces BMI in the primary models, with all other model terms retained. Coefficients involving body mass or linear HKA are reported per 1-SD difference in the corresponding variable; the time x $HKA^2$ coefficient is reported per squared standardized HKA unit. The time x $HKA^2$ terms remained negative and significant in the lateral femur ($\beta = -0.0032$ mm/year, 95% CI [−0.0037, −0.0026], $p < 0.001$) and lateral tibia ($\beta = -0.0032$, 95% CI [−0.0039, −0.0026], $p < 0.001$). Unlike BMI, greater body mass was associated with faster lateral thinning (time x body mass: femur $\beta = -0.0032$, 95% CI [−0.0041, −0.0023]; tibia $\beta = -0.0021$, 95% CI [−0.0032, −0.0009]; both $p < 0.001$). Thus, the lateral estimates ($HKA^2$ x time) were retained, while the association with body size depended on whether BMI or body mass was used.

| OAI Visit | Time From Enrollment | Limbs, n | % | Participants, n |
|---|---|---|---|---|
| V01 | 12 months | 2,424 | 40.9% | 1,263 |
| V03 | 24 months | 2,052 | 34.6% | 1,070 |
| V05 | 36 months | 1,253 | 21.1% | 755 |
| V06 | 48 months | 201 | 3.4% | 104 |
| Total | | 5,930 | 100% | 3,191 |

**Supplementary Table 13. Distribution of the baseline visit relative to the OAI Enrollment.** Baseline was defined for each limb as its first eligible visit with a full-limb alignment (HKA) measurement, complete baseline covariates, and MRI, rather than the enrollment visit (V00). Age, BMI, alignment, and KL grade were taken at each limb's baseline visit, and follow-up time was measured forward from that visit. Most limbs entered at V01 (40.9%) or V03 (34.6%). A participant's two limbs could enter at different visits, so the participant counts across rows exceed the 3,191 unique participants shown in the total.

| Medial Femur (mm) | Underweight (<18.5) | Healthy (18.5–24.9) | Overweight (25.0–29.9) | Obese (≥30) |
|---|---|---|---|---|
| Severe Varus (< −8.8°) | — | 1.03 ± 0.69 | 1.17 ± 0.82 | 1.29 ± 0.71 |
| Moderate Varus (−8.8 to −6.2°) | — | 1.68 ± 0.66 | 1.83 ± 0.61 | 1.58 ± 0.71 |
| Mild Varus (−6.2 to −3.6°) | 1.21 | 1.90 ± 0.49 | 1.90 ± 0.55 | 1.84 ± 0.55 |
| Neutral (−3.6 to 1.6°) | 1.76 ± 0.30 | 1.92 ± 0.39 | 1.99 ± 0.42 | 1.96 ± 0.44 |
| Mild Valgus (1.6 to 4.2°) | 1.83 ± 0.49 | 1.91 ± 0.37 | 2.01 ± 0.38 | 1.96 ± 0.38 |
| Moderate Valgus (4.2 to 6.8°) | 1.25 | 1.91 ± 0.43 | 2.12 ± 0.44 | 2.05 ± 0.33 |
| Severe Valgus (> 6.8°) | — | 2.07 ± 0.56 | 2.17 ± 0.48 | 2.12 ± 0.46 |
| **Medial Tibia (mm)** | **Underweight (<18.5)** | **Healthy (18.5–24.9)** | **Overweight (25.0–29.9)** | **Obese (≥30)** |
| Severe Varus (< −8.8°) | — | 1.41 ± 0.46 | 1.52 ± 0.55 | 1.65 ± 0.52 |
| Moderate Varus (−8.8 to −6.2°) | — | 1.77 ± 0.42 | 1.89 ± 0.46 | 1.79 ± 0.49 |
| Mild Varus (−6.2 to −3.6°) | 1.82 | 1.86 ± 0.36 | 1.91 ± 0.40 | 1.89 ± 0.39 |
| Neutral (−3.6 to 1.6°) | 1.76 ± 0.25 | 1.85 ± 0.31 | 1.91 ± 0.34 | 1.93 ± 0.35 |
| Mild Valgus (1.6 to 4.2°) | 1.35 ± 0.19 | 1.83 ± 0.35 | 1.92 ± 0.35 | 1.89 ± 0.33 |
| Moderate Valgus (4.2 to 6.8°) | 1.00 | 1.85 ± 0.33 | 2.03 ± 0.35 | 1.94 ± 0.36 |
| Severe Valgus (> 6.8°) | — | 1.79 ± 0.49 | 2.00 ± 0.46 | 2.00 ± 0.40 |
| **Lateral Femur (mm)** | **Underweight (<18.5)** | **Healthy (18.5–24.9)** | **Overweight (25.0–29.9)** | **Obese (≥30)** |
| Severe Varus (< −8.8°) | — | 2.08 ± 0.39 | 2.09 ± 0.35 | 2.11 ± 0.54 |
| Moderate Varus (−8.8 to −6.2°) | — | 2.04 ± 0.39 | 2.03 ± 0.35 | 2.06 ± 0.36 |
| Mild Varus (−6.2 to −3.6°) | 2.51 | 1.94 ± 0.32 | 2.03 ± 0.35 | 2.00 ± 0.36 |
| Neutral (−3.6 to 1.6°) | 1.96 ± 0.31 | 1.90 ± 0.33 | 2.01 ± 0.36 | 2.02 ± 0.35 |
| Mild Valgus (1.6 to 4.2°) | 1.47 ± 0.03 | 1.95 ± 0.35 | 2.00 ± 0.33 | 2.02 ± 0.32 |
| Moderate Valgus (4.2 to 6.8°) | 1.50 | 1.91 ± 0.34 | 2.03 ± 0.48 | 2.00 ± 0.41 |
| Severe Valgus (> 6.8°) | — | 1.72 ± 0.51 | 1.77 ± 0.65 | 1.97 ± 0.44 |
| **Lateral Tibia (mm)** | **Underweight (<18.5)** | **Healthy (18.5–24.9)** | **Overweight (25.0–29.9)** | **Obese (≥30)** |
| Severe Varus (< −8.8°) | — | 2.31 ± 0.43 | 2.36 ± 0.49 | 2.44 ± 0.53 |
| Moderate Varus (−8.8 to −6.2°) | — | 2.51 ± 0.52 | 2.53 ± 0.54 | 2.45 ± 0.46 |
| Mild Varus (−6.2 to −3.6°) | 2.57 | 2.42 ± 0.47 | 2.50 ± 0.51 | 2.46 ± 0.52 |
| Neutral (−3.6 to 1.6°) | 2.26 ± 0.46 | 2.38 ± 0.48 | 2.44 ± 0.51 | 2.44 ± 0.52 |
| Mild Valgus (1.6 to 4.2°) | 1.68 ± 0.21 | 2.29 ± 0.58 | 2.27 ± 0.57 | 2.28 ± 0.58 |
| Moderate Valgus (4.2 to 6.8°) | 1.49 | 1.96 ± 0.66 | 2.11 ± 0.70 | 1.98 ± 0.77 |
| Severe Valgus (> 6.8°) | — | 1.34 ± 0.70 | 1.40 ± 0.76 | 1.78 ± 0.76 |

**Supplementary Table 14A. Baseline Cartilage Thickness by Alignment and BMI.** Mean ± SD baseline cartilage thickness (mm) is shown for each region across the alignment and BMI categories used in main manuscript Table 1 (5,930 limbs from 3,191 participants). Limb counts for each alignment and BMI combination are provided in main manuscript Table 1; region summaries use available thickness measurements. Cells with one measurement show the value without an SD; cells with no measurements are indicated by a dash. Negative HKA values indicate varus and positive values indicate valgus. These descriptive summaries provide baseline structural context for the longitudinal analyses.

| **Medial JSN Grade** | **Underweight (<18.5)** | **Healthy (18.5–24.9)** | **Overweight (25.0–29.9)** | **Obese (≥30)** |
|---|---|---|---|---|
| Severe Varus (< −8.8°) | — | 2.40 ± 0.78 | 2.38 ± 0.94 | 2.28 ± 0.81 |
| Moderate Varus (−8.8 to −6.2°) | — | 1.39 ± 1.17 | 1.32 ± 1.04 | 1.71 ± 0.99 |
| Mild Varus (−6.2 to −3.6°) | 2.00 | 0.61 ± 0.90 | 0.96 ± 0.99 | 1.06 ± 1.02 |
| Neutral (−3.6 to 1.6°) | 0.64 ± 1.12 | 0.31 ± 0.60 | 0.46 ± 0.71 | 0.61 ± 0.79 |
| Mild Valgus (1.6 to 4.2°) | 0.00 ± 0.00 | 0.19 ± 0.48 | 0.20 ± 0.47 | 0.39 ± 0.62 |
| Moderate Valgus (4.2 to 6.8°) | 0.00 | 0.07 ± 0.35 | 0.08 ± 0.28 | 0.20 ± 0.50 |
| Severe Valgus (> 6.8°) | — | 0.07 ± 0.27 | 0.18 ± 0.52 | 0.11 ± 0.32 |
| **Lateral JSN Grade** | **Underweight (<18.5)** | **Healthy (18.5–24.9)** | **Overweight (25.0–29.9)** | **Obese (≥30)** |
| Severe Varus (< −8.8°) | — | 0.00 ± 0.00 | 0.00 ± 0.00 | 0.00 ± 0.00 |
| Moderate Varus (−8.8 to −6.2°) | — | 0.02 ± 0.14 | 0.03 ± 0.28 | 0.02 ± 0.15 |
| Mild Varus (−6.2 to −3.6°) | 0.00 | 0.03 ± 0.20 | 0.03 ± 0.22 | 0.03 ± 0.24 |
| Neutral (−3.6 to 1.6°) | 0.36 ± 0.67 | 0.07 ± 0.33 | 0.09 ± 0.40 | 0.11 ± 0.42 |
| Mild Valgus (1.6 to 4.2°) | 0.00 ± 0.00 | 0.27 ± 0.66 | 0.36 ± 0.76 | 0.37 ± 0.77 |
| Moderate Valgus (4.2 to 6.8°) | 0.00 | 0.89 ± 1.13 | 0.82 ± 1.05 | 1.10 ± 1.17 |
| Severe Valgus (> 6.8°) | — | 1.97 ± 1.22 | 2.14 ± 0.98 | 1.61 ± 1.20 |

**Supplementary Table 14B. Baseline Radiographic Joint-Space Narrowing Grades by Alignment and BMI.** Mean ± SD baseline medial and lateral semiquantitative joint-space-narrowing (JSN) grades are shown across the alignment and BMI categories used in main manuscript Table 1. JSN measures were available for all 5,930 limbs from 3,191 participants. Limb counts for each alignment and BMI combination are provided in main manuscript Table 1. Cells with one measurement show the value without an SD; cells with no measurements are indicated by a dash. Negative HKA values indicate varus and positive values indicate valgus. These summaries describe baseline compartment-specific radiographic severity across alignment and BMI groups.

| Term | β [95% CI] | Adjusted OR [95% CI] | p Value |
|---|---|---|---|
| **5 Years: 5,507 Limbs; 190 TKRs** | | | |
| Intercept | -5.6419 [-6.3684, -4.9154] | — | <0.001 |
| HKA Deviation (OR per degree) | 0.2199 [0.0856, 0.3542] | 1.11 [1.04, 1.18] | 0.001 |
| BMI (OR per kg/m²) | 0.1261 [-0.0353, 0.2875] | 1.03 [0.99, 1.06] | 0.126 |
| Age (OR per year) | 0.1502 [-0.0348, 0.3353] | 1.02 [1.00, 1.04] | 0.111 |
| Female vs Male | 0.4479 [0.0765, 0.8193] | 1.57 [1.08, 2.27] | 0.018 |
| KL 1 vs 0 | 0.4770 [-0.5464, 1.5005] | 1.61 [0.58, 4.48] | 0.361 |
| KL 2 vs 0 | 1.7307 [0.9358, 2.5256] | 5.64 [2.55, 12.50] | <0.001 |
| KL 3 vs 0 | 2.6897 [1.9111, 3.4683] | 14.73 [6.76, 32.08] | <0.001 |
| KL 4 vs 0 | 4.0771 [3.2600, 4.8942] | 58.97 [26.05, 133.51] | <0.001 |
| **10 Years: 4,155 Limbs; 405 TKRs** | | | |
| Intercept | -4.5168 [-5.0702, -3.9633] | — | <0.001 |
| HKA Deviation (OR per degree) | 0.2862 [0.1739, 0.3985] | 1.14 [1.08, 1.21] | <0.001 |
| BMI (OR per kg/m²) | 0.1139 [-0.0263, 0.2541] | 1.02 [0.99, 1.05] | 0.111 |
| Age (OR per year) | 0.1689 [0.0243, 0.3136] | 1.02 [1.00, 1.04] | 0.022 |
| Female vs Male | 0.1885 [-0.1010, 0.4780] | 1.21 [0.90, 1.61] | 0.202 |
| KL 1 vs 0 | 1.0920 [0.4337, 1.7503] | 2.98 [1.54, 5.76] | 0.001 |
| KL 2 vs 0 | 1.9081 [1.3135, 2.5027] | 6.74 [3.72, 12.22] | <0.001 |
| KL 3 vs 0 | 3.2330 [2.6609, 3.8050] | 25.35 [14.31, 44.93] | <0.001 |
| KL 4 vs 0 | 4.4763 [3.8379, 5.1147] | 87.91 [46.43, 166.45] | <0.001 |

**Supplementary Table 15. Main-Effects Logistic Models of Recorded Total Knee Replacement Within Five and Ten Years.** Models include absolute HKA deviation from the mean alignment of limbs with KL grade 0 at every matched analytic visit (displayed as −1.0°, with the unrounded mean used in calculations), BMI, age, sex, and categorical baseline KL grade. β coefficients for continuous predictors are per 1-SD difference, using full-cohort baseline standardization. Adjusted odds ratios (ORs) are per degree of HKA deviation, kg/m² of BMI, or year of age. Confidence intervals and p values use participant-clustered robust standard errors. Models were refitted without the HKA deviation x BMI interaction after it was nonsignificant at both horizons (five years, p = 0.361, ten years, p = 0.286). Twenty limbs with self-reported replacement without confirmation were excluded from both analyses. Among the remaining limbs, 403 at five years and 1,755 at ten years were excluded because follow-up ended before the cutoff without recorded TKR. The final analyses included 5,507 limbs with 190 TKRs at five years and 4,155 limbs with 405 TKRs at ten years. Greater HKA deviation was associated with higher TKR odds at both horizons. BMI was not significantly associated with TKR odds.
